# EARLY-ALS: A Multicentre Study on Presymptomatic and Prodromal Amyotrophic Lateral Sclerosis

**DOI:** 10.64898/2026.01.30.26345123

**Authors:** Isabell Cordts, Ana Galhoz, Laura Tzeplaeff, Anne Gründel, Florian Kohlmayer, Andreas Schwersenz, Ines Marschalkowski, Bogdan Bjelica, Verena Panitz, Christian Schulze, Martin Svačina, Petra Rau, André C. Dorigan, Marcus Deschauer, Simon Witzel, David Brenner, Jochen H. Weishaupt, Sarah K. Bublitz, Stefan Lorenzl, Andreas Hermann, Tim Hagenacker, Joachim Wolf, Martin Regensburger, Helmar C. Lehmann, Maike F. Dohrn, Johannes Dorst, René Guenther, André Maier, Thomas Meyer, Daniel Zeller, Christoph Neuwirth, Markus Weber, Jan C. Koch, Matthias Boentert, Markus Weiler, Susanne Petri, Ute Weyen, Torsten Grehl, Michael P. Menden, Paul Lingor

**Affiliations:** Department of Neurology, TUM University Hospital – Klinikum rechts der Isar, TUM School of Medicine, Technical University of Munich, Munich 81675, Germany; Computational Health Center, Helmholtz Munich, Neuherberg 85764, Germany; Bitcare GmbH, Munich 81675, Germany; TUM ForTe, Munich 80333, Germany; Center for ALS and other Motor Neuron Disorders, Department of Neurology, Alfried Krupp Krankenhaus, Essen 45131, Germany; Department of Neurology, Hannover Medical School, Hannover 30625, Germany; PRACTIS Clinician Scientist Program, Dean’s Office for Academic Career Development, Hannover Medical School, Hannover 30625, Germany; Department of Neurology, Heidelberg University Hospital, and Faculty of Medicine, Heidelberg University, Heidelberg 69120, Germany; Department of Neurology, Medical Faculty of the RWTH Aachen University, Aachen 52074, Germany; Department of Neurology, Faculty of Medicine and University Hospital Cologne, University of Cologne, Cologne 50937, Germany; Philipps University Marburg & Department of Neurology, University Hospital Gießen and Marburg, Marburg 35043, Germany; Department of Neurology, University of Ulm, Ulm 89081, Germany; Division of Neurodegeneration, Department of Neurology, Mannheim Center for Translational Neurosciences, Medical Faculty Mannheim, Heidelberg University, Mannheim 68167, Germany; Department of Neurology and Palliative Care, Agatharied Hospital, Hausham 83734, Germany; Institute of Palliative Care, Paracelsus Medical University, Salzburg 5020, Austria; Translational Neurodegeneration Section “Albrecht Kossel”, and Center for Transdisciplinary Neurosciences Rostock (CTNR), University Medical Center Rostock, Rostock 18147, Germany; German Center for Neurodegenerative Diseases (DZNE), Rostock/Greifswald, Rostock 18147, Germany; Department of Neurology, Center for Translational Neuro- and Behavioral Sciences (C-TNBS), University Medicine Essen, Essen 45147, Germany; Department of Neurology, Diako, BruLderklinikum Julia Lanz, Mannheim 68163, Germany; Department of Molecular Neurology, FAU Erlangen-Nürnberg, Erlangen 91054, Germany; Department of Neurology, Faculty of Medicine and University Hospital Carl Gustav Carus, TUD Dresden University of Technology, Dresden 01307, Germany; Charité - Universätsmedizin Berlin, Berlin 10117, Germany; Department of Neurology, University Hospital Würzburg, Würzburg 97080, Germany; HOCH, Kantonsspital St. Gallen, St. Gallen 9007, Switzerland; Department of Neurology, University Medical Center Göttingen, Göttingen 37075, Germany; University of Münster, Münster 48149, Germany; Neuromedical Centre, Department of Sleep Medicine, Klinikum Osnabrück, Osnabrück 49074, Germany; BG University Hospital Bochum, Bochum 44789, Germany; Department of Biochemistry and Pharmacology, Bio21 Molecular Science and Biotechnology Institute, The University of Melbourne, Parkville VIC 3052, Australia; German Center for Neurodegenerative Diseases (DZNE), Site Munich, Munich 81377, Germany; Munich Cluster for Systems Neurology (SyNergy), Munich 81377, Germany

**Keywords:** motor neuron disease, early disease detection, preclinical neurodegeneration, environmental risk factors, sex-specific differences, cohort study

## Abstract

Neurodegenerative diseases often feature a prolonged presymptomatic phase during which pathological processes evolve before overt clinical manifestation. In Amyotrophic lateral sclerosis (ALS), defining this prodromal period is critical for identifying early disease features and the optimal window for intervention, yet it remains poorly characterised.

In this cross-sectional study, we compared 475 ALS patients with 285 controls recruiting across 20 ALS expert centres in Germany and Switzerland. Participants completed a structured digital questionnaire capturing prodromal complaints, healthcare utilisation, comorbidities, lifestyle factors, and weight changes during the 10 years preceding ALS symptom onset.

ALS patients reported substantially higher burden of prodromal complaints than controls (OR=7.50, 95% CI 4.27-13.17; *P* < 0.001; *P*_adj_ < 0.001), particularly neuro-motor, sensory, and pain-related symptoms. Prior to symptom onset, ALS patients more frequently consulted neurologists (OR=1.26, CI 1.10-1.44; *P* < 0.001; *P*_adj_ = 0.007). Speech therapy consultations were significantly more common among female patients (OR = 2.35, CI 1.05–5.28; *P* = 0.038) and those with bulbar-onset ALS (OR = 8.67, CI 3.80–19.77; *P* < 0.001).

Prodromal musculoskeletal dysfunction was more frequently reported by ALS patients and exhibited sex- and site-specific patterns. Herniated discs were reported more often by male ALS patients (OR=2.21, CI 1.04-4.68; *P* = 0.038) and by those with spinal-onset disease (OR=2.32, CI 1.38-3.93; *P* = 0.002). ALS patients more often completed lower secondary education (OR=1.93, CI 1.24-3.01; *P* = 0.004; *P*_adj_ = 0.020) and were more likely to have worked in physically demanding occupations (OR=2.21, CI 1.42-3.43; *P* < 0.001; *P*_adj_ = 0.003).

Lifestyle factors differed significantly, with higher prior consumption of caffeine (OR=7.21, CI 3.27–15.89; *P* < 0.001; *P*_adj_ < 0.001), alcohol (OR=2.25, CI 1.47–3.43; *P* < 0.001; *P*_adj_ = 0.002), and cigarettes (OR=1.64, CI 1.20–2.24; *P* = 0.002) among ALS patients (*P*_adj_ = 0.011). Weight trajectories differed by sex (*P* = 0.009), with male ALS patients showing significant loss already during the pre-onset phase (*P* < 0.001).

These findings demonstrate that ALS is preceded by a distinct prodromal phase characterised by mild motor impairment, altered healthcare engagement, and sex- and site-specific patterns in comorbidities, lifestyle, and metabolic change. Characterising these early features of ALS may facilitate earlier diagnosis and enable timely enrolment in clinical trials.

## Introduction

Neurodegenerative diseases typically evolve through a presymptomatic phase before clinical diagnosis. This phase begins with a clinically silent period in which biomarkers may indicate disease presence (premanifest stage), followed by early symptoms that signal progression but remain insufficient for a definitive clinical diagnosis (prodromal stage). It ultimately transitions to a clinically manifest stage defined by characteristic symptoms permitting clinical diagnosis.^1^ Identifying these early stages is critical for understanding disease mechanisms and for enabling therapeutic and preventive interventions at a time when neuronal damage may still be limited.

Presymptomatic and prodromal phases are well recognised in several neurodegenerative disorders. In Parkinson’s disease, prodromal features such as REM sleep behaviour disorder may precede motor symptoms by years and strongly predict disease.^2^ In Alzheimer’s disease, mild cognitive impairment defines the prodromal stage,^3^ while in Huntington’s disease, subtle motor, cognitive, or psychiatric changes can occur long before clinical diagnosis.^4,5^ In contrast, prodromal markers in amyotrophic lateral sclerosis (ALS) remain incompletely characterised.

Evidence available mostly from genetic cases indicates that ALS is preceded by measurable prodromal changes. Early motor neuron dysfunction in the absence of disease-defining weakness has been reported, particularly in carriers of pathogenic genetic variants, and includes muscle cramps, reduced exercise tolerance, fasciculations, hyperreflexia, and electromyographic abnormalities, collectively referred to as mild motor impairment (MMI).^6^ Subtle cognitive and/or behavioural changes that do not meet criteria for overt frontotemporal dementia (FTD), consistent with mild cognitive and/or mild behavioural impairment (MCI/MBI), may also precede overt muscle atrophy and weakness.^7,8^ Prodromal weight loss is increasingly recognised as an early feature of ALS, is associated with poorer prognosis^9–11^ and hypothalamic atrophy even in presymptomatic carriers of pathogenic genetic variants.^12^ Despite these observations, non-motor symptoms remain poorly studied, underdiagnosed, and underreported in ALS, despite their substantial impact on quality of life, particularly during the prodromal stages. ^13^

Environmental and lifestyle factors are likely to contribute to ALS onset and progression in addition to intrinsic disease mechanisms. As most ALS cases are sporadic and genetic variants explain only part of disease heritability, cumulative lifetime environmental exposures, collectively termed the exposome, are likely to play an important role. In line with the gene-time-environment hypothesis, interaction between genetic susceptibility, aging, and environmental exposures are thought to shape disease onset and progression.^14^ Understanding the ALS exposome is therefore critical as a potentially modifiable component of disease risk.

A wide range of environmental factors have been investigated in ALS, including occupational and environmental exposures (e.g., heavy metals, pesticides, viruses), lifestyle-related factors (e.g., smoking, physical activity, dietary habits), and comorbid medical conditions (e.g., metabolic disorders, cancer, neuroinflammation), yet most reported associations lack consistent or convincing evidence.^15,16^ A meta-analysis identified five environmental factors including heavy metals, pesticides, physical trauma, electric shock, and organic solvents as being associated with ALS onset.^17^ Nevertheless, research in this field remains challenging, as environmental exposures are heterogeneous, often subtle, and accumulate over decades, making them difficult to quantify and posing substantial methodological limitations for epidemiological studies.^18^

In this multicentric study, we aimed to systematically assess self-reported symptoms, environmental exposures, and life events in the pre-manifest phase of ALS. By comparing ALS patients with controls largely drawn from the same environment, we enabled an integrated evaluation of individual symptoms alongside environmental and lifestyle factors within a single study framework.

## Materials and methods

### Patient recruitment

This cross-sectional, multi-centre study was conducted between September 2020 and March 2024 using an online questionnaire. Participants were aged ≥ 18 years and had a diagnosis of possible, probable or definite ALS according to the revised El Escorial criteria.^19^ Patients were recruited at 20 specialized ALS clinics in Germany and Switzerland, mostly affiliated with the Network for Motor Neuron Diseases (MND-NET).^20^ To minimize selection bias, all patients meeting the inclusion criteria and attending the ALS clinics during the recruitment period were identified as potential participants. Proxy-assisted completion was permitted for patients, who were not physically able to fill out the questionnaire themselves.

Participants with another neurodegenerative disease, a neuroinflammatory disorder, or dementia were excluded. Controls consisted primarily of family members, partners, friends, or caregivers recruited either during study visits or via patient invitation. In the same setting, the questionnaire was also administered to patients with motoneuron diseases other than ALS, including spinal muscular atrophy (SMA), hereditary spastic paraplegia (HSP) and spinal and bulbar muscular atrophy (SBMA); analyses of these cohorts will be reported elsewhere. The study was approved by the ethics committees of all participating sites and the lead ethics committee at TUM University Hospital (383/20 S). Written informed consent was obtained from all participants.

### Data collection

The digital questionnaire was developed and conducted using the open-source survey software tool LimeSurvey (https://www.limesurvey.org). Participants accessed the survey via a secure HTTPS connection using unique personal registration tokens provided during routine clinical visits. Both the questionnaire and completed responses were hosted on a secure web server, and patient questionnaires were matched to controls using these tokens. The digital platform enabled a personalised survey flow, with follow-up questions displayed conditionally based on previous responses. The survey comprised seven sections: 1) demographics; 2) disease-related aspects including self-reported ALS phenotype, genetics, and medication; 3) prodromal complaints and early non-motor changes across organ systems; 4) healthcare system interactions; 5) comorbidities; 6) lifestyle changes including occupation, physical activity, diet, and social life; and 7) the self-assessed ALS Functional Rating Scale-Revised (ALSFRS-R). Participants were instructed to skip questions that were not applicable or not relevant to their individual situation.

Most questions referred to the 10-year period prior to ALS symptom onset. Patients were explicitly instructed to consider the time before their first ALS-related symptom, such as muscle weakness, gait disturbance, or swallowing difficulties, and to report changes not previously attributed to ALS or another known medical condition. For controls, the reference period was defined as the 10 years preceding survey completion. In both groups, this period was subdivided into four intervals: 5–10 years, 1–5 years, 1–12 months, and less than 1 month.

Most questions were closed-ended, with predefined response options, and several included an “Other” option allowing free-text input. One open-ended question in the prodromal complaints section invited participants to describe any notable changes in the 10-year reference period that could not be explained by known comorbidities. For these responses and for the comorbidities section, participants indicated the timing of each complaint, which was subsequently assigned to predefined time intervals to ensure consistent temporal categorisation. Responses reporting symptom or condition onset after ALS symptom onset were excluded.

### Data and statistical analyses

Data collected via LimeSurvey were exported to CSV format for analysis and comprised continuous, categorical, binary, and date-time variables. Date-time variables were converted into continuous measures by calculating time in years relative to the questionnaire’s reference date. Missing values in binary variables were imputed to mitigate data sparsity, and continuous variables were normalized to account for scaling differences. Age data were missing for a substantial proportion of participants due to a data capture error affecting this variable alone; to avoid bias or loss of power, age was not included as a covariate in subsequent analyses.

Univariate comparisons between ALS patients and controls were performed using logistic regression, with statistical significance assessed using Wald test. All analyses were stratified and adjusted by sex. Given the self-administrated design and sparsity, analyses were conducted within an exploratory analytical framework, using a conventional p-value (*P*) threshold of *P* <J0.05. Effect sizes were reported as odds ratios (OR) with 95% confidence intervals (CI). False discovery rate (FDR) was controlled using the Benjamini–Hochberg method, and adjusted p-values (*P*_adj_) were reported when significance (*P* < 0.05) was observed. Demographic and clinical group differences were additionally assessed using t-tests or Mann–Whitney tests for continuous variables, depending on distribution and sample size, and Fisher’s exact test for categorical variables.

Subgroup analyses were restricted to questionnaire items significantly enriched in ALS patients in the primary statistical analyses and with sufficient data completeness. Analyses were stratified by sex and site of disease onset. Sex-specific effects were assessed using separate logistic regression models for males and females. Effects by site of onset were evaluated using multinomial logistic regression with a three-level outcome variable comprising control, bulbar-onset ALS and spinal-onset ALS, followed by pairwise contrasts. Subgroup results were reported as OR with 95% CI and associated p-values.

Weight trajectories were analysed using linear mixed-effects models (LMMs) with time as a categorical variable and participant-specific random intercepts. Models included group (ALS vs. controls), sex, time, and their interactions to assess disease- and sex-specific effects.

## Results

### Demographics and characteristics of the study groups

Demographic and clinical characteristics of the study groups in the EARLY-ALS multicentre case–control study were derived from 818 enrolled participants, including 513 ALS patients and 305 controls; 58 participants (38 ALS and 20 controls) only partially completed the questionnaires and were excluded, resulting in final analytic cohort of 760 participants, comprising 475 ALS patients and 285 controls.

The cohort captures a broad demographic and clinical landscape of ALS spanning disease onset, phenotype, functional status, and genetic background (Table 1). In the ALS group, there were significantly more males. Significant differences in sex were observed between ALS patients and controls (*P* < 0.001), with no age difference between groups. The ALS symptom onset was reported with a median of 2 years (IQR: 1-4 years) before questionnaire completion. Most ALS patients reported a “classical ALS” phenotype (69.47%). A genetic cause of disease was reported by approximately 3.58% of ALS patients.

### Questionnaire overview

Both the ALS and control group completed a core questionnaire covering seven categories. Including all main, sub-, and conditional items, 558 questions were presented, with ALS patients answering an additional 52 disease-specific questions (Supplementary Tables 1 and 2). Excluding free-text responses yielded 491 questions for univariate analysis (Supplementary Fig. 1A), comprising 196 continuous, 191 binary, and 104 categorical variables. As non-applicable items were left blank by design, missing values in binary variables and questions concerning doctor visits were imputed as zero (Supplementary Fig. 1B).

### Prodromal complaints and early non-motor changes

The ALS cohort reported significantly more prodromal changes prior to symptom onset compared with controls (OR=7.50, CI 4.27–13.17; *P* < 0.001; *P*_adj_ < 0.001). After exclusion of non-informative free-text entries, 117 ALS patients (24.63%) and 15 controls (5.26%) reported 261 and 25 prodromal changes, respectively. ALS patients who reported prodromal changes, 61.54% described two or more changes and 38.46% reported only one. Reported changes were manually categorised into 13 distinct symptom domains (Fig. 1A).

**Figure 1:**
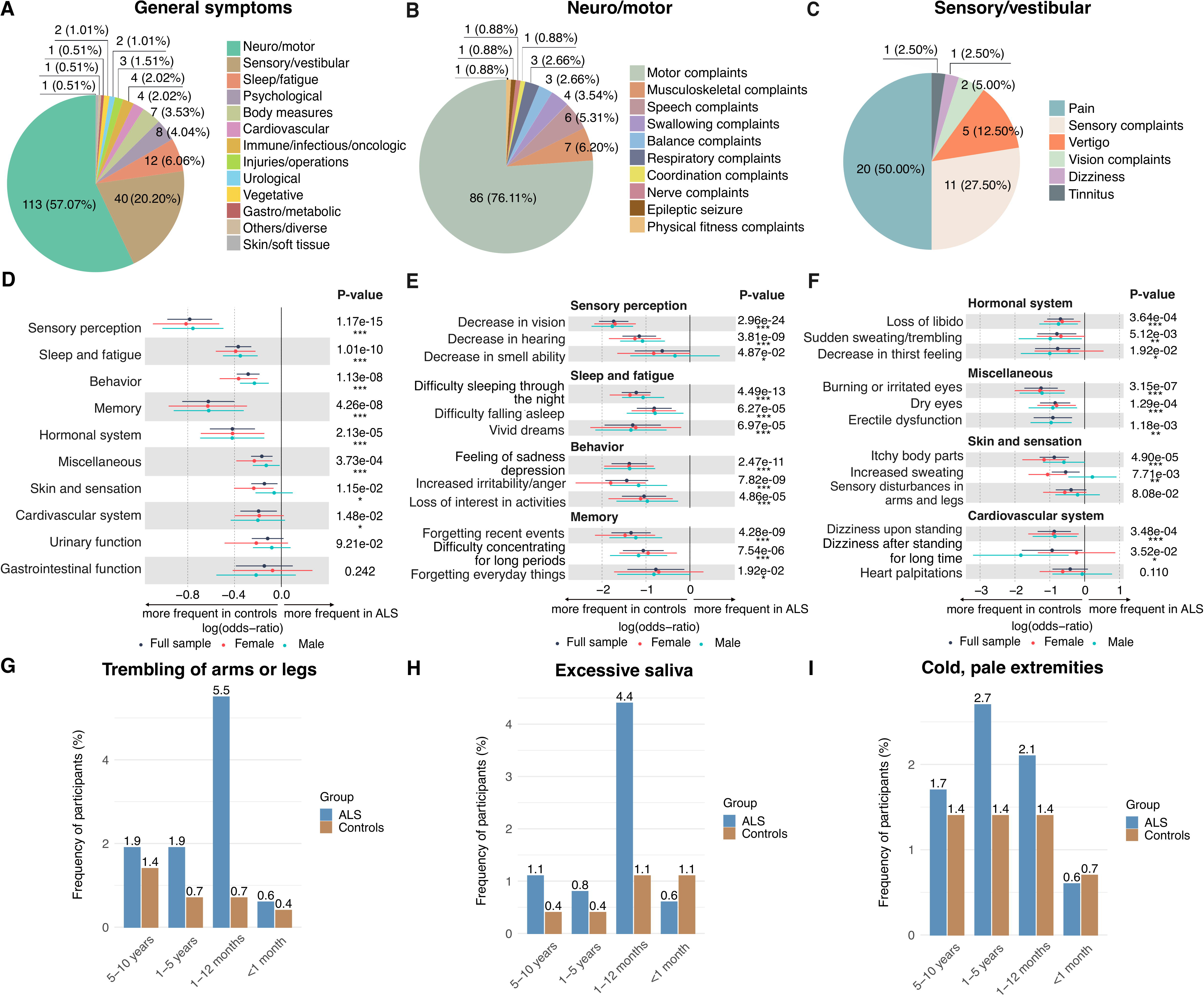
Prodromal complaints of ALS patients and control participants. **(A)-(C)** Prodromal complaints provided by ALS patients in response to an open-ended question. **(A)** Pie chart showing the relative frequencies of the 13 symptom domains into which the open-ended responses were manually categorized. Subcategories within the most frequently reported domains, **(B)** neuro/motor domain and **(C)** sensory/vestibular domain are visualized. **(D)** Forest plot for ten predefined categories of non-motor complaints reported by ALS patients and healthy individuals. **(E)-(F)** Forest plots of the eight significant prodromal complaints (*P <* 0.05) and their top three subcategories (based on lowest p-values). These reflect results from univariate analyses based on the full sample, i.e., all participants adjusted for sex (black), and sex-stratified analyses (female in salmon, male in blue). The p-values available in the forest plots are specific to the full sample analysis, and significance is indicated as *P <* 0.05 (*), *P <* 0.01 (**), *P <* 0.001 (***). **(G)–(I)** Bar plots displaying the frequency of the three prodromal symptoms reported more frequently by ALS patients than controls, originating from the miscellaneous and skin/sensation categories: **(G)** trembling of arms or legs, **(H)** excessive saliva, and **(I)** cold, pale extremities. The percentage of participants reporting each symptom is shown for four time intervals: 5–10 years, 1–5 years, 1–12 months, and <1 month before symptom onset (ALS patients) or questionnaire completion (controls).

Prodromal changes reported by ALS patients were dominated by neuro/motor (57.07%) and sensory/vestibular (20.20%) symptoms (Fig. 1B-C). Within the neuro/motor domain, motor complaints predominated (76.11%), most commonly muscle weakness, followed by muscle cramps and gait disturbances. In the sensory/vestibular category, pain was most frequent (50.00%), particularly back pain, followed by muscle and limb pain. Notably, both neuro/motor and sensory/vestibular symptoms were commonly reported to have been present several years before ALS onset (defined based on symptoms in Supplementary Table 3), with median intervals of 1 year (IQR: 0-3) and 2 years (IQR: 1-7), respectively (Supplementary Fig. 2).

Non-motor prodromal symptoms were reported more frequently by controls than by ALS patients. Using 78 targeted questions across ten symptom categories (Fig. 1D), 40 symptoms were significantly more common in controls (OR<1; *P* < 0.05), whereas only one was enriched in ALS patients (OR>1; *P* < 0.05; Fig. 1D–F). Symptoms less frequently reported by ALS patients clustered predominantly in the domains of sensory perception, sleep and fatigue, and behaviour; the most frequently reported symptoms within each of these domains are shown in Fig. 1E. Only few non-motor symptoms were enriched in ALS patients and clustered close to symptom onset. Trembling of the arms or legs was the most frequent prodromal complaint (OR=3.24, CI 1.55-6.78; *P* = 0.002; *P*_adj_ = 0.011), while we observed a trend for excessive saliva (OR=2.22, CI 1.00-4.95; *P* = 0.051), and cold, pale extremities (OR=1.58, CI 0.82-3.03; *P* = 0.173). These symptoms were most commonly reported within 1–12 months before ALS onset (Fig. 1G-I).

### Healthcare interactions

Prodromal healthcare utilisation in ALS was characterised by increased neurology consultations. Over the 10 years preceding ALS symptom onset or survey completion, ALS patients reported more frequent visits to neurologists than controls (OR=1.26, CI 1.10-1.44; *P* < 0.001; *P*_adj_ = 0.007; Fig. 2A), with most visits occurring withing 1-12 months before symptom onset (Fig. 2B). Visits to speech therapists were also more common in ALS patients, particularly in women, but did not reach statistical significance when considering the full cohort (OR=1.11, CI 0.99-1.25; *P* = 0.076; Fig. 2C). In contrast, controls more frequently consulted with specialists in oncology (OR=0.89, CI 0.82-0.97; *P* = 0.011; *P*_adj_ = 0.048), dermatology (OR=0.93, CI 0.87-0.99; *P* = 0.027; *P*_adj_ = 0.100) and endocrinology (OR=0.92, CI 0.85-0.99; *P* = 0.032; *P*_adj_ = 0.116) (Fig. 2A).

**Figure 2:**
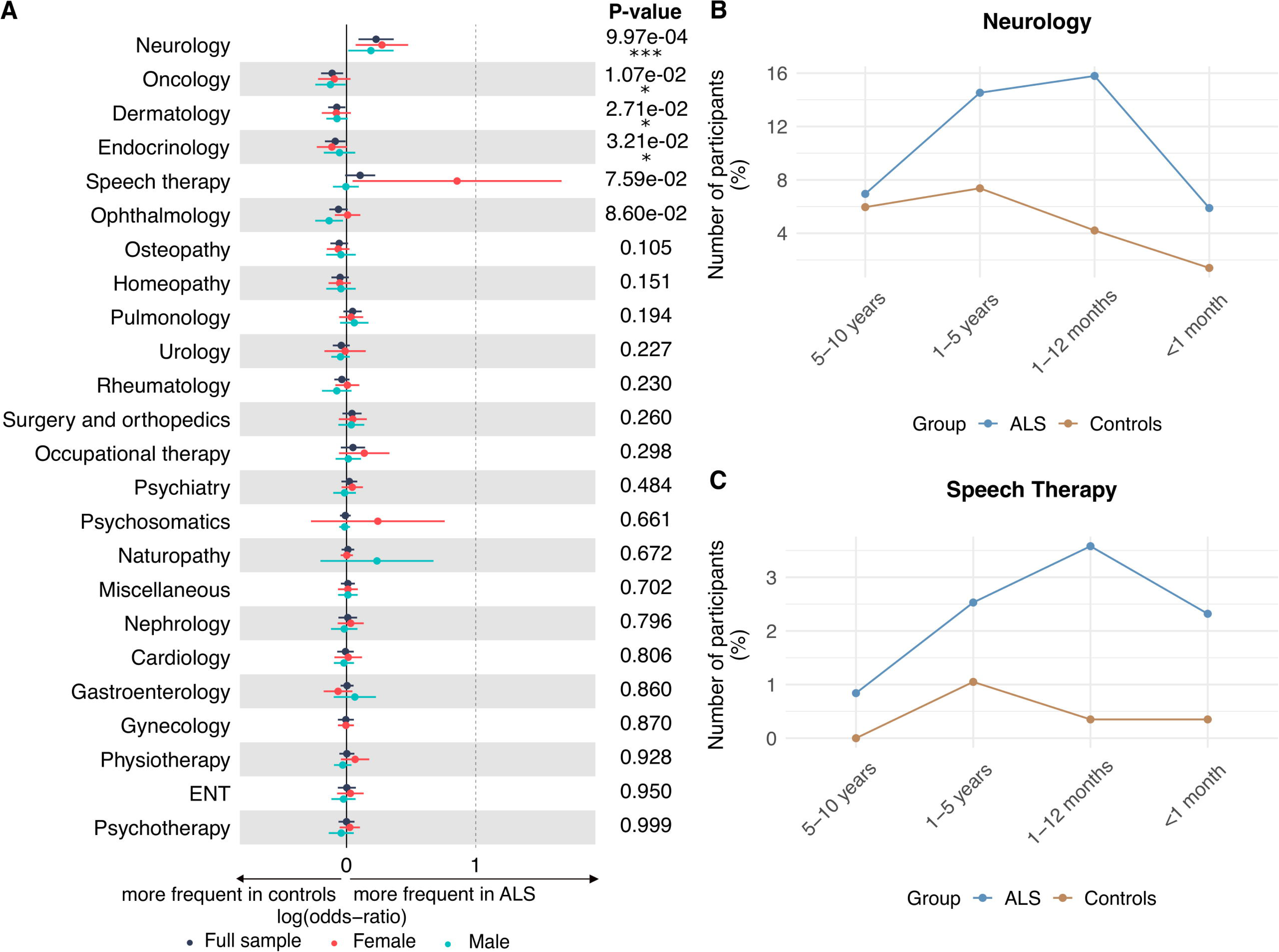
Healthcare interactions of ALS patients and control participants. **(A)** Frequency of interactions with 24 different medical specialists reported by ALS patients and controls. Forest plot illustrates univariate results based on the full sample, i.e., all participants adjusted for sex (black), and sex-stratified analyses (female in salmon, male in blue). The p-values are specific to the full sample analysis and significance is indicated as *P <* 0.05 (*), *P <* 0.01 (**), *P <* 0.001 (***). **(B)-(C)** Percentage of patients who consulted neurologists and speech therapists, respectively, at four time intervals prior to symptom onset (ALS patients) or questionnaire completion (controls). Percentages represent the number of participants who consulted each specialist during a specific period divided by the total number of participants.

### Comorbidities

Musculoskeletal comorbidities were more frequently reported in ALS patients than in controls. Pre-existing musculoskeletal disorders prior to ALS symptom onset were more common in the ALS group (OR=1.46, CI 1.04-2.05; *P* = 0.029; *P*_adj_ = 0.108; Fig. 3A). Within this category, herniated disc was the only condition reaching statistical significance, reported by 13.89% of ALS patients compared with 7.37% of controls (OR=1.85, CI 1.10-3.13; *P* = 0.021; *P*_adj_ = 0.086; Fig. 3B). Of the ALS patients reporting herniated disc, 40 (60.61%) provided timing information. Most events (67.50%) occurred 5–10 years prior to ALS symptom onset (Fig. 3C).

**Figure 3:**
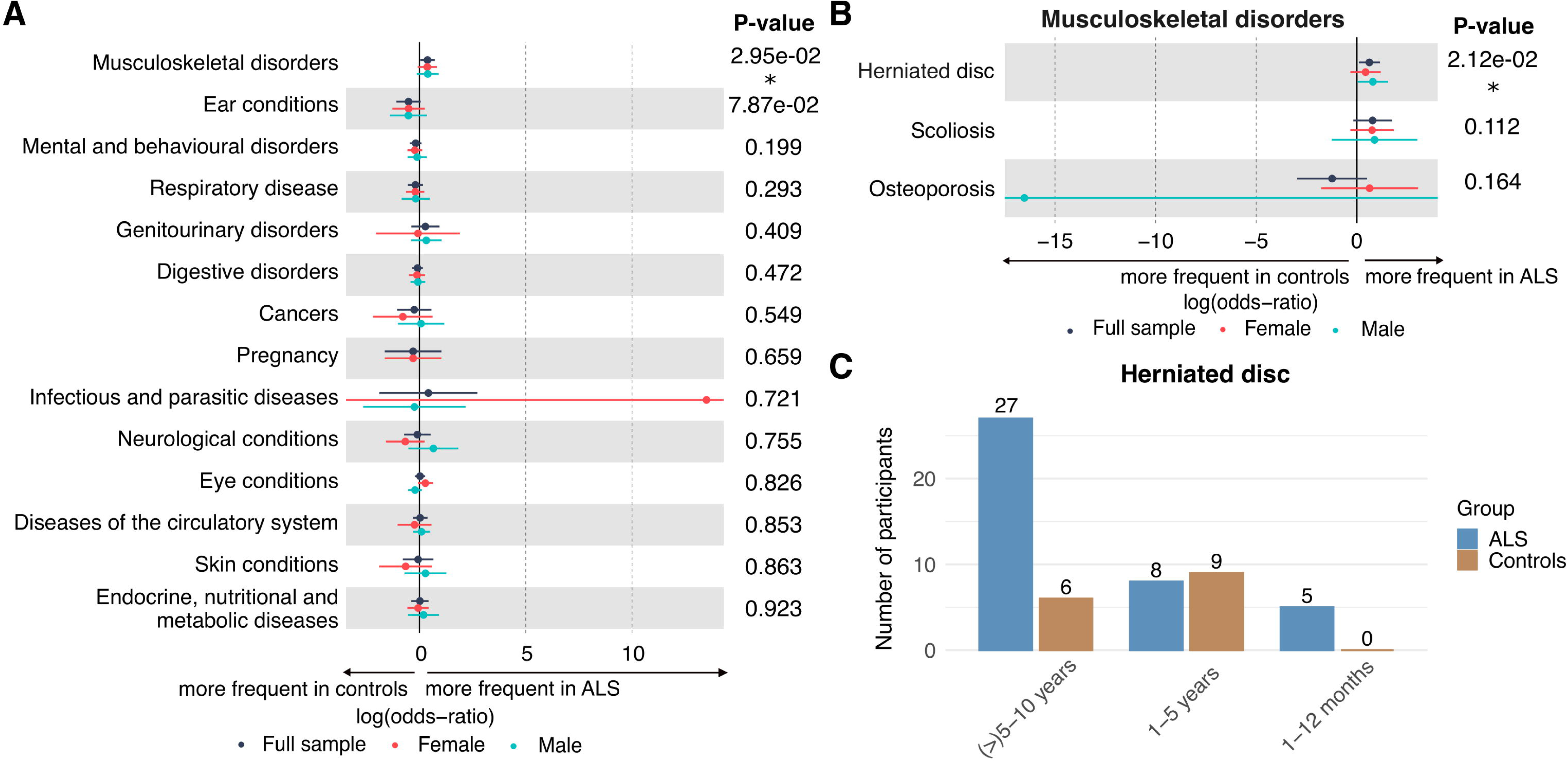
Pre-existing comorbidities in ALS patients and control participants. **(A)** Forest plot of univariate analyses comparing the frequency of 14 main categories of comorbidities between ALS patients and healthy controls. While the original questionnaire assessed 15 comorbidity categories, those related to miscellaneous conditions were answered using free-text and were therefore excluded from this plot. Results reflect univariate analyses based on the full sample, i.e., all participants adjusted for sex (black), and sex-stratified analyses (female in salmon, male in blue). P-values correspond to the full sample analysis and significance is indicated as *P <* 0.05 (*). **(B)** Visualization of top three specific conditions within the musculoskeletal disorders category. Results are from univariate analyses based on the full sample, i.e., all participants adjusted for sex (black), and sex-stratified analyses (female in salmon, male in blue). P-values correspond to the full sample analysis and significance is indicated as *P <* 0.01 (**). **(C)** Distribution of herniated disc cases across time intervals: (>)5-10 years, 1-5 years and 1-12 months prior to symptom onset (ALS patients) or prior to questionnaire completion (controls). Participants reported the exact date of onset for herniated disc diagnoses. For consistency, these dates were manually grouped into time intervals aligned with those used in other questions. Cases reported more than 10 years before symptom onset or questionnaire completion were included in the “(>)5–10 years” interval. Specifically, 22 of the 27 responses (ALS) and 5 of the 6 responses (controls) assigned to this interval referred to events occurring more than 10 years before symptom onset or questionnaire completion.

### Lifestyle changes and sociobiographical events

Social and life-event profiles differed between ALS patients and controls. ALS patients were more likely to report being single (OR=2.07, CI 1.26-3.39; *P* = 0.004; *P*_adj_ = 0.022), and to have children (OR=1.41, CI 1.01-1.98; *P* = 0.045; *P*_adj_ = 0.155; Fig. 4A). In contrast, when major life events were considered collectively, controls reported significantly more personal and professional changes (OR=0.31, CI 0.22-0.43; *P* < 0.001; *P*_adj_ < 0.001), driven largely by reports of serious illness affecting a close person (OR=0.13, CI 0.09-0.18; *P* < 0.001; *P*_adj_ < 0.001), consistent with the predominance of relatives among controls (Fig. 4A, Table 1). Other major life events did not differ between groups (Supplementary Table 4). Infection-related experiences differed minimally between groups: only common colds were reported more frequently by controls (OR=0.45, CI 0.30-0.67; *P* < 0.001; *P*_adj_ = 0.001) (Fig. 4A).

**Figure 4:**
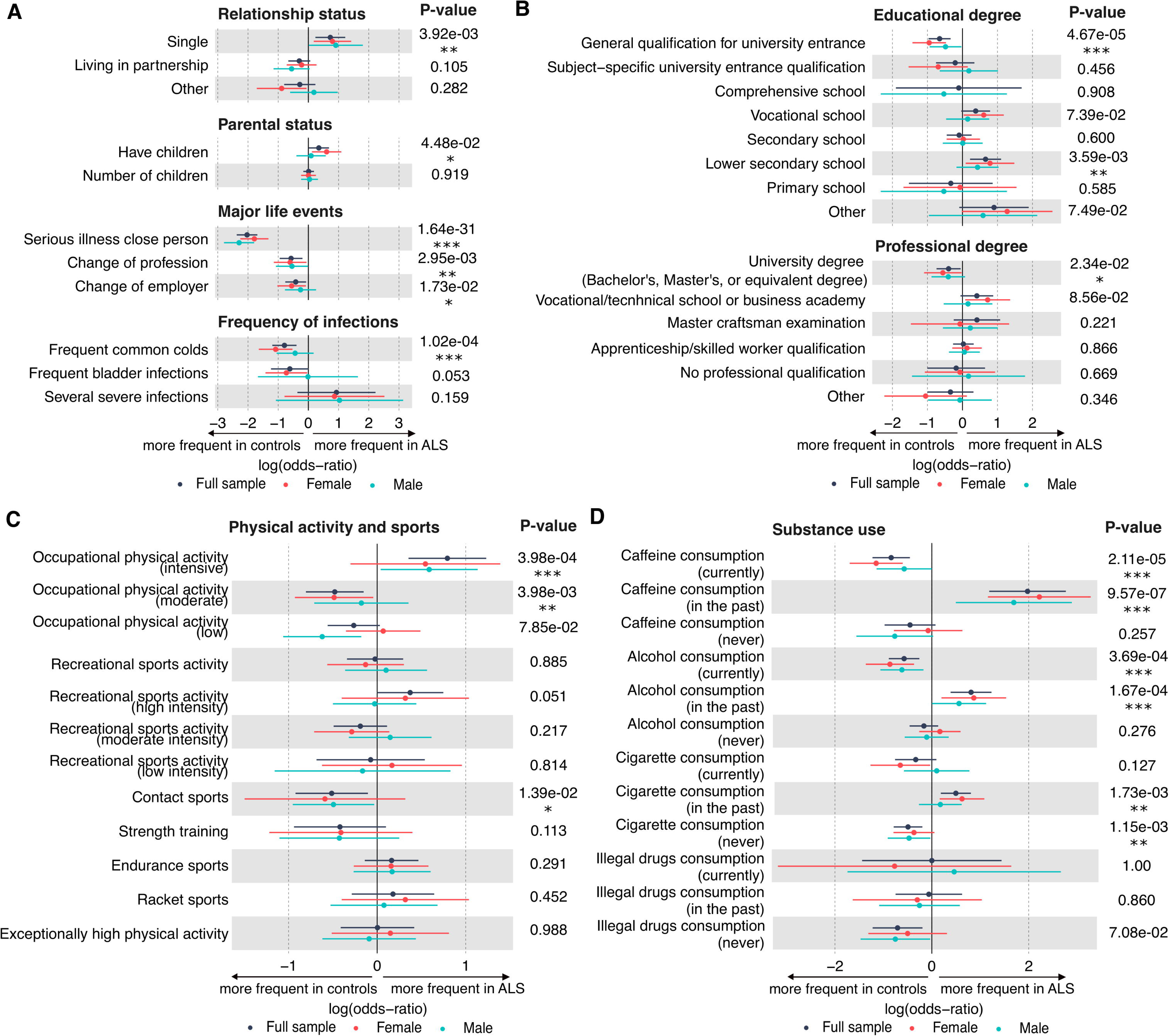
Lifestyle differences between ALS patients and control participants. **(A)** Forest plots illustrating differences in relationship and parental status, major life events, and infection frequency. For life events and infections, only the top three subcategories (based on the lowest p-values) are displayed. **(B)** Distribution of educational and professional qualifications, presented in descending order where applicable. **(C)** Differences in physical activity, including work-related physical exertion (categorized from intensive to low), recreational engagement in sports, types of sports practiced and the occurrence of episodes of exceptionally high physical activity (e.g., running a marathon). **(D)** Patterns of substance use, including caffeine, alcohol, and cigarette consumption, categorized by current use, past use, or never. All panels show univariate analyses results for the full sample adjusted for sex (black), and sex-stratified analyses (female in salmon, male in blue). P-values are specific to the full sample analysis, and significance is indicated as *P <* 0.05 (*), *P <* 0.01 (**), *P <* 0.001 (***).

Educational attainment differed between ALS patients and controls. ALS patients more frequently reported completion of lower secondary school (OR=1.93, CI 1.24–3.01; *P* = 0.004; *P*_adj_ = 0.020), whereas controls more often reported holding a general qualification for university entrance (OR=0.52, CI 0.38–0.71; *P* < 0.001; *P*_adj_ < 0.001) and university degrees (OR=0.67, CI 0.48–0.95; *P* = 0.023; *P*_adj_ = 0.089) (Fig. 4B).

Occupational but not recreational physical activity differed between ALS patients and controls. ALS patients were more likely to engage in physically demanding occupations (OR=2.21, CI 1.42-3.43; *P* < 0.001; *P*_adj_ = 0.003), whereas controls more frequently reported jobs involving moderate physical activity (OR=0.62, CI 0.45-0.86; *P* = 0.004; *P*_adj_ = 0.022) (Fig. 4C). In contrast, no group differences were observed in regular sports participation. The distribution of intensity of recreational activity or the frequency of exceptionally intense physical exertion within the 10 years preceding ALS symptom onset or survey completion was not different. Only contact sports were reported more frequently by controls, although this did not reach adjusted significance (OR=0.60, CI 0.40–0.90; *P* = 0.014; *P*_adj_ = 0.060) (Fig.4C).

Substance use in ALS was characterised by higher past but lower current consumption. ALS patients more frequently reported past consumption of caffeinated drinks (OR=7.21, CI 3.27-15.89; *P* < 0.001; *P*_adj_ < 0.001), alcohol (OR=2.25, CI: 1.47-3.43; *P* < 0.001; *P*_adj_ = 0.002), and cigarettes or other tobacco products (OR=1.64, CI 1.20-2.24; *P* = 0.002; *P*_adj_ = 0.011) (Fig. 4D). In contrast, controls were more likely to report current use of caffeine (OR=0.43, CI 0.29-0.64; *P* < 0.001; *P*_adj_ < 0.001) and alcohol (OR=0.56, CI 0.41-0.77; *P* < 0.001; *P*_adj_ = 0.003), and to have never smoked (OR=0.61, CI: 0.45-0.82; *P* = 0.001; *P*_adj_ = 0.008) (Fig. 4D; Supplementary Fig. 3). Quantities of substances used were not significantly different between groups (Supplementary Fig. 4).

### Diet and weight

Dietary patterns were largely comparable between groups, apart from calorie-restricted diets. Over the 10 years preceding symptom onset or survey completion, controls more frequently reported following a calorie-restricted diet (OR=0.58, CI 0.39-0.85; *P* = 0.006; *P*_adj_ = 0.029). No significant differences were observed for fasting, adopting a vegetarian or vegan diet, or other dietary modifications (Supplementary Table 4).

Weight loss emerged before symptom onset and progressed over time in ALS patients. At questionnaire completion, ALS patients weighed significantly less than controls (OR=0.63, CI 0.53–0.75; P < 0.001; *P*_adj_ < 0.001), corresponding to an average difference of 6.4kg (Fig. 5A). While no significant group differences were observed at earlier time points, controls generally maintained stable or slightly increased weight over the preceding 10 years. In contrast, ALS patients reported weight loss beginning 1–12 months prior to symptom onset, with mean loss of 5.6kg from the year before onset to study completion. Additional weight loss was reported between symptom onset and diagnosis (1kg), and from diagnosis to study completion (3.4kg).

**Figure 5:**
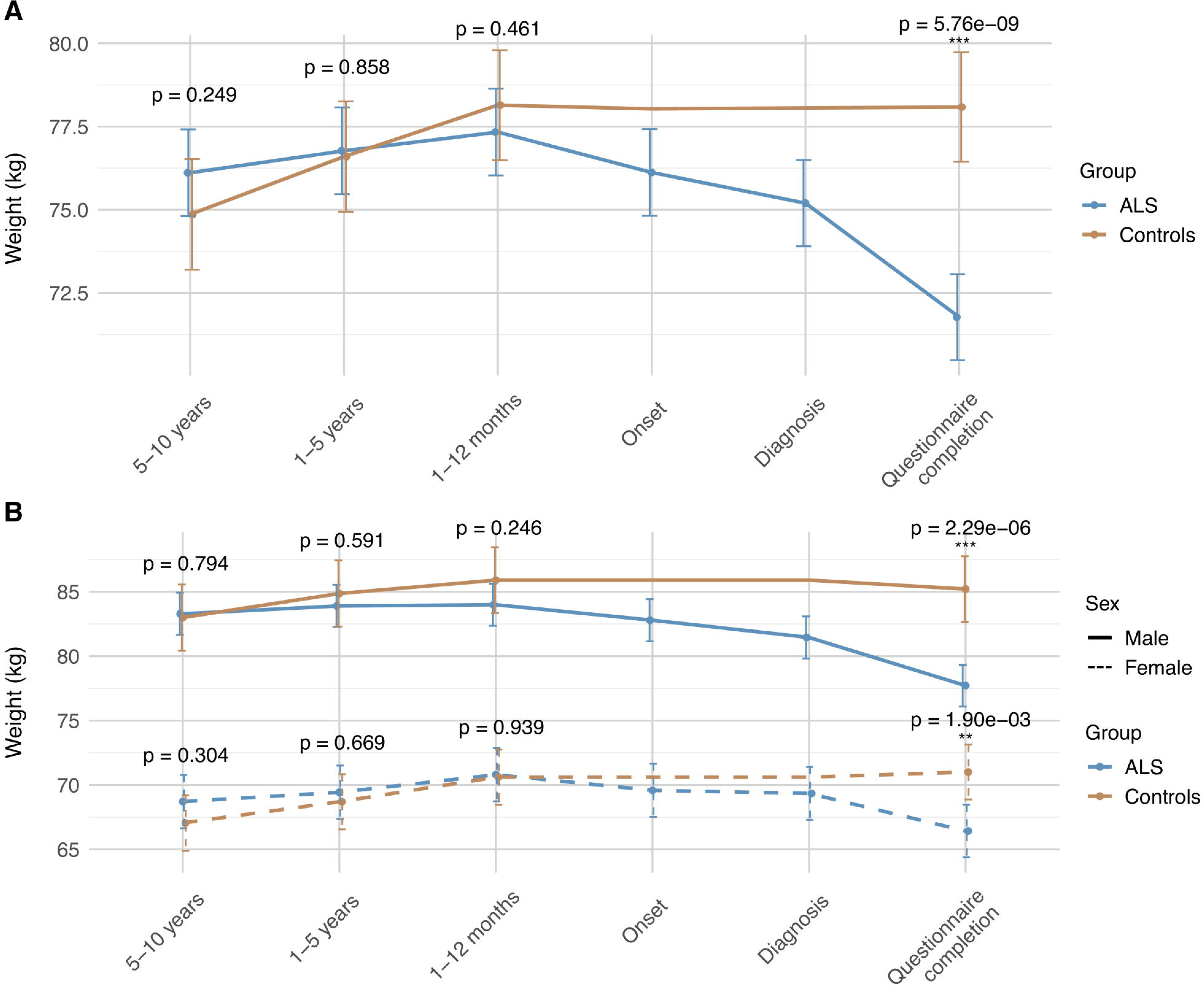
Retrospective weight trajectories in ALS patients and healthy controls. **(A)** Weight data for ALS patients and controls across six retrospective time periods, 5–10 years, 1–5 years, and 1–12 months before symptom onset (ALS) or time of questionnaire completion (controls), as well as at symptom onset (ALS), at diagnosis (ALS), and at questionnaire completion (both groups). Weight trajectories are shown as marginal means adjusted for sex, estimated from a linear mixed-effect model (LMM) with patient as a random effect and fixed effects for group, sex and time. **(B)** Weight data for ALS patients and controls stratified by sex, across the same time periods as in **(A)**. Trajectories were obtained from an LMM with patient as a random effect and fixed effects of time and group. Solid lines represent male participants and dashed lines represent female participants. In both panels, lines indicate mean values with 95% confidence intervals. Statistical significance between groups was assessed using pairwise contrasts from the LMM, and is indicated as *P <* 0.05 (*), *P <* 0.01 (**), *P <* 0.001 (***).

### Subgroup analyses

Exploratory subgroup analyses revealed sex-specific patterns in prodromal non-motor symptoms in ALS (Supplementary Tables 5 and 6). While trembling of the arms and legs was more frequently reported by both female (OR=3.18, CI 1.14–8.88; *P* = 0.027) and male ALS patients (OR=3.30, CI 1.14–9.58; *P* = 0.028) compared with their respective controls, excessive saliva was only reported more often by female ALS patients (OR=10.87, CI 1.39–85.06; *P* = 0.023).

Subgroup analyses by site of disease onset identified distinct prodromal symptom profiles (Supplementary Table 7). Trembling of arms and legs was more frequently reported only by spinal-onset ALS patients compared with controls (OR=3.43, CI 1.63–7.21; *P* = 0.001). In contrast, bulbar-onset patients were significantly more likely to report excessive saliva than both spinal-onset patients (OR=2.92, CI 1.34–6.35; *P* = 0.007) and controls (OR=5.94, CI 2.30–15.37; *P* < 0.001).

Healthcare utilisation prior to ALS symptom onset varied by sex and site of disease onset. Neurology visits were more frequently reported in both female (OR=1.31, CI 1.07–1.61; *P* = 0.008) and male ALS patients (OR=1.21, CI 1.01–1.44; *P* = 0.036) compared with controls, and were increased in both bulbar-onset (OR=3.55, CI 2.03–6.23; *P* < 0.001) and spinal-onset ALS (OR=2.93, CI 2.00–4.29; *P* < 0.001). Speech therapy visits were only more commonly reported by female ALS patients (OR=2.35, CI 1.05–5.28; *P* = 0.038) and by bulbar-onset patients compared with both controls (OR=8.67, CI 3.80–19.80; *P* < 0.001) and spinal-onset patients (OR=3.66, CI 1.92–7.00; p < 0.001), while spinal-onset patients differed only from controls (OR=2.37, CI 1.14–5.93; *P* = 0.021).

Herniated disc was the only pre-existing condition showing sex- and site-specific associations in ALS. In subgroup analyses, herniated disc was more frequently reported by male ALS patients (OR=2.21, CI 1.04–4.68; *P* = 0.038) and by spinal-onset patients (OR=2.32, CI 1.38–3.93; *P* = 0.002) compared with controls. Adjustment for physically demanding occupations did not affect these associations, which remained significant in both male ALS patients (OR=2.73, CI 1.16-7.52; *P* = 0.032) and spinal-onset ALS patients (OR=2.34, CI 1.32-4.33; *P* = 0.005).

Sex- and site-of-onset–specific differences shaped social, educational, occupational, and lifestyle characteristics in ALS. Consistent with univariate analyses, ALS patients were more often single than controls, including women (OR=2.22, CI 1.19–4.13; *P* = 0.012) and men (OR=2.48, CI 1.01–6.07; *P* = 0.047), and spinal- (OR=1.81, CI 1.08–3.04; *P* = 0.024) and bulbar-onset ALS (OR=2.91, CI 1.43–5.93; *P* = 0.003). Female ALS patients were more likely to have children compared to controls than males (OR=1.83, CI 1.13–2.97; *P* = 0.015), and this was also reported by spinal-onset patients (OR=1.64, CI 1.15–2.33; *P* = 0.006). Educational differences were driven by females (Supplementary Fig. 5), who more frequently reported lower secondary school (OR=2.20, CI 1.10–4.40; *P* = 0.026), and vocational or technical qualifications (OR=2.05, CI 1.08–3.90; *P* = 0.028), and spinal-onset ALS patients, who more often completed lower secondary education (OR=1.96, CI 1.23–3.10; *P* = 0.004) (Supplementary Fig. 5).

Occupational and lifestyle factors also differed by sex and site of onset. Male ALS patients were more likely to report physically demanding occupations (OR=1.80, CI 1.04–3.11; *P* = 0.035), with the strongest effect observed in spinal-onset ALS compared with controls (OR=2.59, CI 1.64–4.07; *P* < 0.001) (Supplementary Fig. 6).

Past substance use showed marked sex-specific patterns: among females, past caffeine (OR=9.20, CI 3.18–26.60; *P* < 0.001), alcohol (OR=2.38, CI 1.22–4.64; *P* = 0.011), and smoking (OR=1.86, CI 1.18–2.96; *P* = 0.008) were more frequent in ALS, whereas among male ALS patients only caffeine (OR=5.43, CI 1.64–17.99; *P* = 0.005) and alcohol use (OR=1.75, CI 1.00–3.06; *P* = 0.049) were increased. There was no difference in regard to site-of-onset (Supplementary Fig. 7).

Reported weight loss in ALS was sex-dependent, began years before symptom onset, and progressed over time. Across the study period, ALS males reported a mean loss of 5.6kg and ALS females of 2.3kg, whereas control males and females reported a gain of 2.2 kg and 4.0 kg, respectively. Across all presymptomatic time points up to 10 years before onset, male ALS patients consistently reported lower weight than male controls, while no such group difference was observed among females (Fig. 5B). An ALS-only linear mixed-effects model showed a significant time-by-sex interaction during the pre-onset period (*P* = 0.009) (Fig. 5B), and sex-stratified models confirmed greater weight loss only in male ALS patients compared with controls (P < 0.001) (Supplementary Table 8). Reported weight loss continued after symptom onset, with males losing 1.3 kg from onset to diagnosis and a further 3.7 kg thereafter, and females losing 0.3 kg and 2.9 kg, respectively. At questionnaire completion, male ALS patients weighed −7.4 kg relative to controls (OR=0.58, CI 0.45–0.75; *P* < 0.001), and females −4.7 kg (OR=0.69, CI 0.54–0.89; *P* = 0.004).

Site-of-onset analyses revealed early weight differences in spinal-onset ALS. Spinal-onset patients reported lower weight than controls 5-10 years (OR=1.02, CI 1.01–1.03; *P* = 0.002) and 1-5 years (OR=1.01, CI 1.00–1.03; *P* = 0.006) before symptom onset. At questionnaire completion, both bulbar-onset (OR=0.98, CI 0.96–1.00; *P* = 0.016) and spinal-onset ALS (OR= 0.99, CI 0.98–1.00; *P* = 0.025) weighted less than controls (Supplementary Table 7).

## Discussion

While a prolonged prodromal phase is well recognized in other neurodegenerative diseases, the early stages of ALS are less well defined. By the time diagnostic criteria are fulfilled, substantial motor neuron loss has already occurred.^21^ Identifying early disease-related changes is critical to enable earlier diagnosis, improve patient stratification, optimise clinical trial design, and ultimately improve therapeutic outcomes.^1^

In this study, ALS patients reported a markedly higher burden of prodromal symptoms than controls, predominantly affecting the neuromuscular system. Despite explicit instructions to report only symptoms preceding recognised ALS onset, 18.53% of patients described motor-related complaints, most commonly weakness and muscle cramps. Trembling of extremities was the most frequently reported non-motor symptom; however, this likely reflects subtle early motor involvement rather than a true non-motor feature. Together, these findings suggest that patients perceive neuromuscular changes before objective clinical deficits become apparent, indicating very early motor dysfunction in the sense of a mild motor impairment.

Healthcare utilisation increased during the prodromal phase of ALS, suggesting a window for early clinical recognition. ALS patients reported more frequent consultations with neurologists (35.16%) and speech therapists (8.00%) during this period, indicating that patients actively sought medical advice prior to formal diagnosis.

These observations support the concept of mild motor impairment, previously described mainly in presymptomatic carriers of ALS-associated genetic variants, and extend it to a broad, predominantly sporadic ALS cohort.^22^ This is consistent with recent reports showing prodromal, primarily mild motor symptoms occurring within six months before recognised ALS onset in approximately one quarter of patients.^23^

Sensory complaints represented the second most frequent prodromal feature, with pain being the most common. Sensory abnormalities, including paraesthesia, hypoesthesia, and features of sensory neuropathy, have been reported in approximately 20–30% of ALS patients, depending on the method of assessment.^24,25^ Pain can arise at any disease stage, and up to half of patients recall pain or sensory symptoms months to years before motor onset or diagnosis.^26–28^ Recognising pain as a potential prodromal feature is clinically relevant, as sensory symptoms may prompt alternative diagnostic considerations and delay ALS diagnosis and treatment initiation.^29^

We observed a higher incidence of musculoskeletal disorders, particularly herniated discs, prior to ALS symptom onset. This may partly reflect diagnostic bias, as early non-specific neuromuscular complaints often lead to spinal imaging and detection of incidental, but not necessarily causative disc pathology. However, because most herniated discs were reported 5–10 years before ALS onset, simple misattribution of early symptoms seems less likely. Nevertheless, orthopaedic misdiagnosis remains common in ALS, with some patients undergoing inappropriate spinal surgery.^30,31^ Alternatively, disc prolapse itself may contribute to focal neural injury that could initiate or accelerate neurodegeneration.^32^

In contrast to other neurodegenerative disorders, we identified relatively few specific non-motor prodromal symptoms in ALS. This may reflect systematic underreporting, as patients retrospectively prioritise salient motor deficits while subtle non-motor changes fade from memory leading to recall bias. Supporting this interpretation, ALS patients may report normal subjective sleep despite marked alterations in sleep architecture^33^, potentially related to involvement of von Economo neurons in the anterior cingulate cortex, which are critical for subjective judgment.^34^ Moreover, since impaired self-awareness or agnosia is common in frontotemporal dementia (FTD),^35–37^ and (mild) cognitive/behavioural impairments occur in around 50% of ALS patients,^38,39^ reduced awareness of symptoms cannot be excluded.

The elevated burden of non-motor complaints in controls potentially reflects caregiving-related psychosocial stress rather than prodromal disease features. Controls, many of whom were spouses or relatives of ALS patients, reported more sleep and behavioural symptoms, consistent with increased healthcare utilisation outside neurology or higher rates of infections.^40,41^ By contrast, ALS patients, who have been described as exhibiting high agreeableness, ^42^ may be less inclined to modify life circumstances despite emerging difficulties, potentially contributing to lower reporting of such non-motor changes.

ALS patients, particularly females, more frequently reported lower educational attainment, alongside a higher prevalence of physically demanding occupations, especially among males. These findings align prior evidence linking manual labour and lower socioeconomic status to increase ALS risk.^43–45^ In contrast, lifetime engagement in recreational sports was similar between groups, suggesting that occupational exposures, characterised by sustained intensity and cumulative burden, may be more relevant than leisure activity.^46,47^ Potential exposure to environmental toxins, more common in physically demanding occupations, including heavy metals, pesticides, or solvents, also have been implicated in ALS pathogenesis.^45,48^

ALS patients reported higher past but lower current use of caffeine, alcohol, and cigarettes, whereas controls more often report ongoing consumption, likely reflecting lifestyle changes following symptom onset or diagnosis. While overall substance exposure was similar, ALS patients showed a trend toward higher high-level smoking in the past, consistent with epidemiological evidence linking smoking to increased ALS risk.^49,50^ In contrast, evidence for alcohol remains inconsistent. ^51–53^ While caffeine has been suggested to exert neuroprotective effects in Parkinson’s disease even at relatively low intake,^54^ current and prospective studies show no association between pre-diagnostic caffeine intake and ALS risk, ^55^ with experimental data suggesting potential harm in *SOD1* mouse models.^56^

Controls more frequently reported calorie-restricted diets, possibly reflecting a reduced need for dietary restriction in ALS due to disease-related weight loss or behavioural changes affecting lifestyle habits. Consistent with previous studies, ALS patients showed substantial early weight loss,^57,58^ a key non-motor feature associated with poorer prognosis.^11^ This weight loss likely reflects a combination of muscle wasting, reduced intake, and hypermetabolism linked to hypothalamic dysfunction that may precede motor symptoms.^59–61^ In our cohort, weight loss was reported 1–12 months before symptom onset and continued thereafter, underscoring the importance of early and sustained nutritional management in ALS.

Subgroup analyses revealed distinct sex- and site-of-onset-specific patterns, underscoring the value of stratified approaches for identifying early ALS signatures. Excessive saliva was a prominent in female and bulbar-onset patients, and coincided with increased pre-diagnostic speech therapy visits, likely reflecting the higher prevalence of bulbar-onset ALS among females.^62^ Pre-existing herniated discs were more frequent in males and spinal-onset ALS patients. The epidemiology of disc herniation is inconsistent with some surgical cohorts reporting a slight male predominance,^63^ while others describe higher rates in women.^64^ In our cohort, males and spinal-onset patients were also more likely to have held physically demanding jobs. Although lumbar disc herniation has been linked to manual labour,^64,65^ intensive physical work did not explain this association in our cohort, suggesting additional contributions from chronic spinal microtrauma, sex-specific biology, or delayed healthcare-seeking in men.

Consistent with prior reports of greater weight loss during disease progression^66^, we show that pre-morbid weight loss is also disproportionately reported by men. Together with emerging evidence of more pronounced molecular alterations in male ALS patients,^67^ these sex-specific patterns may converge to increase motor neuron vulnerability, providing a potential mechanistic basis for sex differences in ALS prevalence and clinical phenotype.

The retrospective design of this study entails inherent limitations. Recall bias is a major concern, particularly for vague or nonspecific symptoms, and reporting accuracy may be further reduced by mild cognitive impairment in ALS. Reliance on self-reported data limits objectivity, whereas clinical assessments or biomarkers could more reliably characterise prodromal features.^68^ Furthermore, despite the relatively large cohort, statistical power may still be insufficient to detect smaller effects or subtle subgroup differences.

Our findings suggest that ALS is often diagnosed after a prodromal phase during which neurological and/or speech therapy services are already engaged, highlighting missed opportunities for earlier recognition. By establishing the relevance of the prodromal mild motor impairment in a large, multicentric, predominantly sporadic ALS cohort, this study provides a foundation for refining clinical frameworks to identify ALS at earlier stages. Moreover, the observation that weight loss emerges closer to ALS symptom onset supports its inclusion as a candidate early marker for future risk stratification and trial enrichment.

## Data Availability

All data produced in the present study are available upon reasonable request to the authors.

## Acknowledgements

We thank all patients and their caregivers or relatives who participated in this trial. BB was supported by PRACTIS Clinician Scientist Program, funded by Hannover Medical School and DFG (DFG ME 3696/3). MW is a member of the European Reference Network for Neuromuscular Diseases (ERN EURO-NMD).

## Competing interests

AH received grants from Hermann and Lilly Schilling Stiftung für medizinische Forschung im Stifterverband, BMBF/VDI, DFG, consulting fees from Zambon, ITF Pharma, and royalties from Kohlhammer and Elsevier.

TH reports consulting fees from Biogen, Novartis, ScholarRock, Roche, SanofiGenzyme, Alexion, Amgen, Argenx, AstraZeneca, Alnylam, UCB and Santhera and speaker fees from Biogen, Novartis, Roche, SanofiGenzyme, Alexion, Argenx, AstraZeneca, Alnylam and UCB.

MFD received financial reimbursement for consulting and advisory board activities and travel support to attend scientific meetings by Akcea Therapeuticals Inc., Alnylam Pharmaceuticals Inc., Amgen Therapeutics, Amicus Therapeutics, Applied Therapeutics, Astra Zeneca, Pfizer Pharmaceuticals, and Swedish Orphan Biovitrum AB.

MB has received research support from Sanofi and Löwenstein Medical and consulting fees or speaker honoraria from Amicus, Amylyx, Biogen, ITF Pharma, Sanofi, Sentec, and Zambon.

MW reports consultant and advisory board honoraria from Akcea Therapeutics, Alnylam Pharmaceuticals, Biogen, F. Hoffmann-La Roche, Novartis, Novo Nordisk, Pfizer, Purpose Pharma, and Swedish Orphan Biovitrum AB; speaker fees from Akcea Therapeutics, Alnylam Pharmaceuticals, AstraZeneca, and Biogen; and financial support for conference or meeting attendances from Akcea Therapeutics, Alnylam Pharmaceuticals, AstraZeneca, BridgeBio, Ionis, and Pfizer, outside of the scope of the present work.

MPM is a former employee of AstraZeneca (Cambridge, UK). MPM collaborates with and receives financial support from GlaxoSmithKline (London, UK), F. Hoffmann-La Roche (Basel, Switzerland), and AstraZeneca (Cambridge, UK). MPM also provides consultancy services to Merck Sharp & Dohme (MSD) and McKinsey & Company.

PL reports grants from the Bundesministerium für Bildung und Forschung and the Deutsche Forschungsgemeinschaft; consulting fees from AbbVie, Amylyx, Bial, Desitin, ITF Pharma, Novartis, Stadapharm, Raya Therapeutic, Woolsey Pharmaceuticals, Trace Neuroscience and Zambon; and is co-inventor on a patent for the use of fasudil in amyotrophic lateral sclerosis (EP 2825175 B1, US 9.980,972 B2), outside of the scope of the submitted work.

SP received honoraria as a speaker/consultant from Biogen GmbH, Roche, Novartis, Cytokinetics Inc., Desitin, Italfarmaco, Amylyx, and Zambon; and grants from DGM e.V., Federal Ministry of Education and Research, German Israeli Foundation for Scientific Research and Development, EU Joint Program for Neurodegenerative Disease Research, and Neurodegenerative Research Inc., outside of the submitted work.

The other authors report no COI regarding this specific project.

## Supplementary material

Supplementary material is available at *Brain* online.

## References

1. Benatar M, Wuu J, McHutchison C, et al. Preventing amyotrophic lateral sclerosis: insights from pre-symptomatic neurodegenerative diseases. Brain. 2021;

2. Postuma RB, Iranzo A, Hu M, et al. Risk and predictors of dementia and parkinsonism in idiopathic REM sleep behaviour disorder: a multicentre study. Brain. 2019;142(3):744–759.

3. Morris JC, Storandt M, Miller JP, et al. Mild cognitive impairment represents early-stage Alzheimer disease. Archives of neurology. 2001;58(3):397–405.

4. Paulsen J, Langbehn DR, Stout JC, et al. Detection of Huntington’s disease decades before diagnosis: the Predict-HD study. *Journal of Neurology*, Neurosurgery & Psychiatry. 2008;79(8):874–880.

5. Duff K, Paulsen JS, Beglinger LJ, Langbehn DR, Stout JC. Psychiatric symptoms in Huntington’s disease before diagnosis: the predict-HD study. Biol Psychiatry. Dec 15 2007;62(12):1341–6. doi:10.1016/j.biopsych.2006.11.034

6. Benatar M, Granit V, Andersen PM, et al. Mild motor impairment as prodromal state in amyotrophic lateral sclerosis: a new diagnostic entity. Brain. 2022;145(10):3500–3508. doi:10.1093/brain/awac185

7. Strong MJ, Abrahams S, Goldstein LH, et al. Amyotrophic lateral sclerosis - frontotemporal spectrum disorder (ALS-FTSD): Revised diagnostic criteria. Amyotrophic Lateral Sclerosis and Frontotemporal Degeneration. 2017;18(3-4):153–174. doi:10.1080/21678421.2016.1267768

8. Benatar M, Wuu J, Huey ED, et al. The Miami Framework for ALS and related neurodegenerative disorders: an integrated view of phenotype and biology. Nature Reviews Neurology. 2024/06/01 2024;20(6):364–376. doi:10.1038/s41582-024-00961-z

9. O’Reilly ÉJ, Wang H, Weisskopf MG, et al. Premorbid body mass index and risk of amyotrophic lateral sclerosis. Amyotrophic Lateral Sclerosis and Frontotemporal Degeneration. 2013;14(3):205–211.

10. Moglia C, Calvo A, Grassano M, et al. Early weight loss in amyotrophic lateral sclerosis: outcome relevance and clinical correlates in a population-based cohort. *Journal of Neurology*, Neurosurgery & Psychiatry. 2019;90(6):666. doi:10.1136/jnnp-2018-319611

11. van Mantgem MRJ, van Eijk RP, van der Burgh HK, et al. Prognostic value of weight loss in patients with amyotrophic lateral sclerosis: a population-based study. *Journal of Neurology*, Neurosurgery & Psychiatry. 2020;91(8):867–875.

12. Gorges M, Vercruysse P, Müller H-P, et al. Hypothalamic atrophy is related to body mass index and age at onset in amyotrophic lateral sclerosis. J Neurol Neurosurg Psychiatry. 2017;88(12):1033–1041.

13. Bjelica B, Bartels M-B, Hesebeck-Brinckmann J, Petri S. Non-motor symptoms in patients with amyotrophic lateral sclerosis: current state and future directions. Journal of Neurology. 2024;271(7):3953–3977. doi:10.1007/s00415-024-12455-5

14. Goutman SA, Hardiman O, Al-Chalabi A, et al. Emerging insights into the complex genetics and pathophysiology of amyotrophic lateral sclerosis. The Lancet Neurology. 2022;21(5):465–479.

15. Bozzoni V, Pansarasa O, Diamanti L, Nosari G, Cereda C, Ceroni M. Amyotrophic lateral sclerosis and environmental factors. Functional neurology. 2016;31(1):7.

16. Couratier P, Corcia P, Lautrette G, Nicol M, Preux P-M, Marin B. Epidemiology of amyotrophic lateral sclerosis: a review of literature. Revue neurologique. 2016;172(1):37–45.

17. Wang M-D, Little J, Gomes J, Cashman NR, Krewski D. Identification of risk factors associated with onset and progression of amyotrophic lateral sclerosis using systematic review and meta-analysis. Neurotoxicology. 2017;61:101–130.

18. Al-Chalabi A, Hardiman O. The epidemiology of ALS: a conspiracy of genes, environment and time. Nature Reviews Neurology. 2013;9(11):617–628.

19. Brooks BR, Miller RG, Swash M, Munsat TL. El Escorial revisited: revised criteria for the diagnosis of amyotrophic lateral sclerosis. Amyotrophic lateral sclerosis and other motor neuron disorders. 2000;1(5):293–299.

20. German Network for Motor Neuron Diseases (MND-NET). https://mnd-net.de/de/

21. Mitsumoto H, Kasarskis EJ, Simmons Z. Hastening the diagnosis of amyotrophic lateral sclerosis. Neurology. 2022;99(2):60–68.

22. Benatar M, Cai X, McDermott MP, et al. Proposed Research Criteria for Mild Motor Impairment as a Prodromal Syndrome in Amyotrophic Lateral Sclerosis. Neurology. Aug 26 2025;105(4):e213917. doi:10.1212/wnl.0000000000213917

23. van Wijk IF, Kraneburg L, van Eijk RP, et al. Prodromal symptoms in amyotrophic lateral sclerosis from the perspective of the patient and of the caregiver. Amyotrophic Lateral Sclerosis and Frontotemporal Degeneration. 2025:1–11.

24. Hammad M, Silva A, Glass J, Sladky J, Benatar M. Clinical, electrophysiologic, and pathologic evidence for sensory abnormalities in ALS. Neurology. 2007;69(24):2236–2242.

25. Bombaci A, Lupica A, Pozzi FE, Remoli G, Manera U, Di Stefano V. Sensory neuropathy in amyotrophic lateral sclerosis: a systematic review. Journal of neurology. 2023;270(12):5677–5691.

26. Pota V, Sansone P, De Sarno S, et al. Amyotrophic Lateral Sclerosis and Pain: A Narrative Review from Pain Assessment to Therapy. Behav Neurol. 2024;2024:1228194. doi:10.1155/2024/1228194

27. Chiò A, Mora G, Lauria G. Pain in amyotrophic lateral sclerosis. Lancet Neurol. Feb 2017;16(2):144–157. doi:10.1016/s1474-4422(16)30358-1

28. Taga A, Schito P, Trapasso MC, Zinno L, Pavesi G. Pain at the onset of Amyotrophic Lateral Sclerosis: a cross-sectional study. Clin Neurol Neurosurg. Nov 2019;186:105540. doi:10.1016/j.clineuro.2019.105540

29. Bombaci A, Lupica A, Pozzi FE, Remoli G, Manera U, Di Stefano V. Sensory neuropathy in amyotrophic lateral sclerosis: a systematic review. J Neurol. Dec 2023;270(12):5677–5691. doi:10.1007/s00415-023-11954-1

30. Donaghy C, Dick A, Hardiman O, Patterson V. Timeliness of diagnosis in motor neurone disease: a population-based study. The Ulster medical journal. 2008;77(1):18.

31. Yoshor D, Klugh III A, Appel SH, Haverkamp LJ. Incidence and characteristics of spinal decompression surgery after the onset of symptoms of amyotrophic lateral sclerosis. Neurosurgery. 2005;57(5):984–989.

32. Yip PK, Malaspina A. Spinal cord trauma and the molecular point of no return. Molecular neurodegeneration. 2012;7(1):6.

33. Guillot SJ, Lang C, Simonot M, et al. Early-onset sleep alterations found in patients with amyotrophic lateral sclerosis are ameliorated by orexin antagonist in mouse models. Science translational medicine. 2025;17(783):eadm7580.

34. Lulé D, Michels S, Finsel J, et al. Clinicoanatomical substrates of selfish behaviour in amyotrophic lateral sclerosis–an observational cohort study. Cortex. 2022;146:261–270.

35. Goldstein LH, Abrahams S. Changes in cognition and behaviour in amyotrophic lateral sclerosis: nature of impairment and implications for assessment. Lancet Neurol. Apr 2013;12(4):368–80. doi:10.1016/s1474-4422(13)70026-7

36. Scherling CS, Zakrzewski J, Datta S, et al. Mistakes, Too Few to Mention? Impaired Self-conscious Emotional Processing of Errors in the Behavioral Variant of Frontotemporal Dementia. Front Behav Neurosci. 2017;11:189. doi:10.3389/fnbeh.2017.00189

37. Mendez MF, Shapira JS. Loss of emotional insight in behavioral variant frontotemporal dementia or “frontal anosodiaphoria”. Conscious Cogn. Dec 2011;20(4):1690–6. doi:10.1016/j.concog.2011.09.005

38. Jellinger KA. Mild cognitive impairment in amyotrophic lateral sclerosis: Current view. Journal of Neural Transmission. 2025;132(3):357–368.

39. Jellinger KA. The spectrum of behavioral disorders in amyotrophic lateral sclerosis: current view. Journal of Neural Transmission. 2025;132(2):217–236.

40. Stenson K, Chew S, Dong S, Heithoff K, Wang MJ, Rosenfeld J. Health care resource utilization and costs across stages of amyotrophic lateral sclerosis in the United States. J Manag Care Spec Pharm. Nov 2024;30(11):1239–1247. doi:10.18553/jmcp.2024.30.11.1239

41. Schischlevskij P, Cordts I, Günther R, et al. Informal Caregiving in Amyotrophic Lateral Sclerosis (ALS): A High Caregiver Burden and Drastic Consequences on Caregivers’ Lives. Brain Sciences. 2021;11(6):748.

42. Parkin Kullmann JA, Hayes S, Pamphlett R. Are people with amyotrophic lateral sclerosis (ALS) particularly nice? An international online case–control study of the Big Five personality factors. Brain and Behavior. 2018;8(10):e01119.

43. Sutedja NA, Veldink JH, Fischer K, et al. Lifetime occupation, education, smoking, and risk of ALS. Neurology. Oct 9 2007;69(15):1508–14. doi:10.1212/01.wnl.0000277463.87361.8c

44. Farrugia Wismayer M, Borg R, Farrugia Wismayer A, et al. Occupation and amyotrophic lateral sclerosis risk: a case-control study in the isolated island population of Malta. Amyotroph Lateral Scler Frontotemporal Degener. Nov 2021;22(7-8):528–534. doi:10.1080/21678421.2021.1905847

45. Duan QQ, Jiang Z, Su WM, et al. Risk factors of amyotrophic lateral sclerosis: a global meta-summary. Front Neurosci. 2023;17:1177431. doi:10.3389/fnins.2023.1177431

46. Lacorte E, Ferrigno L, Leoncini E, Corbo M, Boccia S, Vanacore N. Physical activity, and physical activity related to sports, leisure and occupational activity as risk factors for ALS: A systematic review. Neuroscience & biobehavioral reviews. 2016;66:61–79.

47. Chapman L, Cooper-Knock J, Shaw PJ. Physical activity as an exogenous risk factor for amyotrophic lateral sclerosis: a review of the evidence. Brain. 2023;146(5):1745–1757.

48. Su FC, Goutman SA, Chernyak S, et al. Association of Environmental Toxins With Amyotrophic Lateral Sclerosis. JAMA Neurol. Jul 1 2016;73(7):803–11. doi:10.1001/jamaneurol.2016.0594

49. Kim K, Ko DS, Kim JW, et al. Association of smoking with amyotrophic lateral sclerosis: A systematic review, meta-analysis, and dose-response analysis. Tob Induc Dis. 2024;22doi:10.18332/tid/175731

50. Wang H, O’Reilly É J, Weisskopf MG, et al. Smoking and risk of amyotrophic lateral sclerosis: a pooled analysis of 5 prospective cohorts. Arch Neurol. Feb 2011;68(2):207–13. doi:10.1001/archneurol.2010.367

51. Westeneng H-J, van Veenhuijzen K, van der Spek RA, et al. Associations between lifestyle and amyotrophic lateral sclerosis stratified by C9orf72 genotype: a longitudinal, population-based, case-control study. The Lancet Neurology. 2021/05/01/ 2021;20(5):373–384. 10.1016/S1474-4422(21)00042-9

52. D’Ovidio F, Rooney JPK, Visser AE, et al. Association between alcohol exposure and the risk of amyotrophic lateral sclerosis in the Euro-MOTOR study. J Neurol Neurosurg Psychiatry. Jan 2019;90(1):11–19. doi:10.1136/jnnp-2018-318559

53. Peng B, Yang Q, R BJ, et al. Role of Alcohol Drinking in Alzheimer’s Disease, Parkinson’s Disease, and Amyotrophic Lateral Sclerosis. Int J Mol Sci. Mar 27 2020;21(7)doi:10.3390/ijms21072316

54. Palacios N, Gao X, McCullough ML, et al. Caffeine and risk of Parkinson’s disease in a large cohort of men and women. Movement Disorders. 2012;27(10):1276–1282.

55. Fondell E, O’Reilly ÉJ, Fitzgerald KC, et al. Intakes of caffeine, coffee and tea and risk of amyotrophic lateral sclerosis: Results from five cohort studies. Amyotrophic Lateral Sclerosis and Frontotemporal Degeneration. 2015;16(5-6):366–371.

56. Potenza RL, Armida M, Ferrante A, et al. Effects of chronic caffeine intake in a mouse model of amyotrophic lateral sclerosis. Journal of neuroscience research. 2013;91(4):585–592.

57. Mariosa D, Beard JD, Umbach DM, et al. Body mass index and amyotrophic lateral sclerosis: a study of US military veterans. American journal of epidemiology. 2017;185(5):362–371.

58. Moglia C, Calvo A, Grassano M, et al. Early weight loss in amyotrophic lateral sclerosis: outcome relevance and clinical correlates in a population-based cohort. *Journal of Neurology*, Neurosurgery & Psychiatry. 2019;90(6):666–673.

59. Bouteloup C, Desport J-C, Clavelou P, et al. Hypermetabolism in ALS patients: an early and persistent phenomenon. Journal of neurology. 2009;256(8):1236–1242.

60. Gorges M, Vercruysse P, Müller H-P, et al. Hypothalamic atrophy is related to body mass index and age at onset in amyotrophic lateral sclerosis. *Journal of Neurology*, Neurosurgery & Psychiatry. 2017;88(12):1033–1041.

61. Peter RS, Rosenbohm A, Dupuis L, et al. Life course body mass index and risk and prognosis of amyotrophic lateral sclerosis: results from the ALS registry Swabia. European journal of epidemiology. 2017;32(10):901–908.

62. McCombe PA, Henderson RD. Effects of gender in amyotrophic lateral sclerosis. Gend Med. Dec 2010;7(6):557–70. doi:10.1016/j.genm.2010.11.010

63. Strömqvist F, Strömqvist B, Jönsson B, Karlsson MK. Gender differences in patients scheduled for lumbar disc herniation surgery: a National Register Study including 15,631 operations. European Spine Journal. 2016;25(1):162–167.

64. Kim Y-K, Kang D, Lee I, Kim S-Y. Differences in the incidence of symptomatic cervical and lumbar disc herniation according to age, sex and national health insurance eligibility: a pilot study on the disease’s association with work. International journal of environmental research and public health. 2018;15(10):2094.

65. Ahsan M, Matin T, Ali M, Ali M, Awwal M, Sakeb N. Relationship between physical work load and lumbar disc herniation. Mymensingh medical journal: MMJ. 2013;22(3):533–540.

66. Grassano M, Moglia C, Palumbo F, et al. Sex differences in amyotrophic lateral sclerosis survival and progression: a multidimensional analysis. Annals of Neurology. 2024;96(1):159–169.

67. Caldi Gomes L, Hänzelmann S, Hausmann F, et al. Multiomic ALS signatures highlight subclusters and sex differences suggesting the MAPK pathway as therapeutic target. Nature communications. 2024;15(1):4893.

68. Tzeplaeff L, Galhoz A, Meijs C, et al. Identification of a presymptomatic and early disease signature for amyotrophic lateral sclerosis (ALS): protocol of the premodiALS study. Neurological research and practice. 2025;7(1):56.

