## Supplementary Figure for "EARLY-ALS: A Multicentre Study on Presymptomatic and Prodromal Amyotrophic Lateral Sclerosis"

### Supplementary Figures

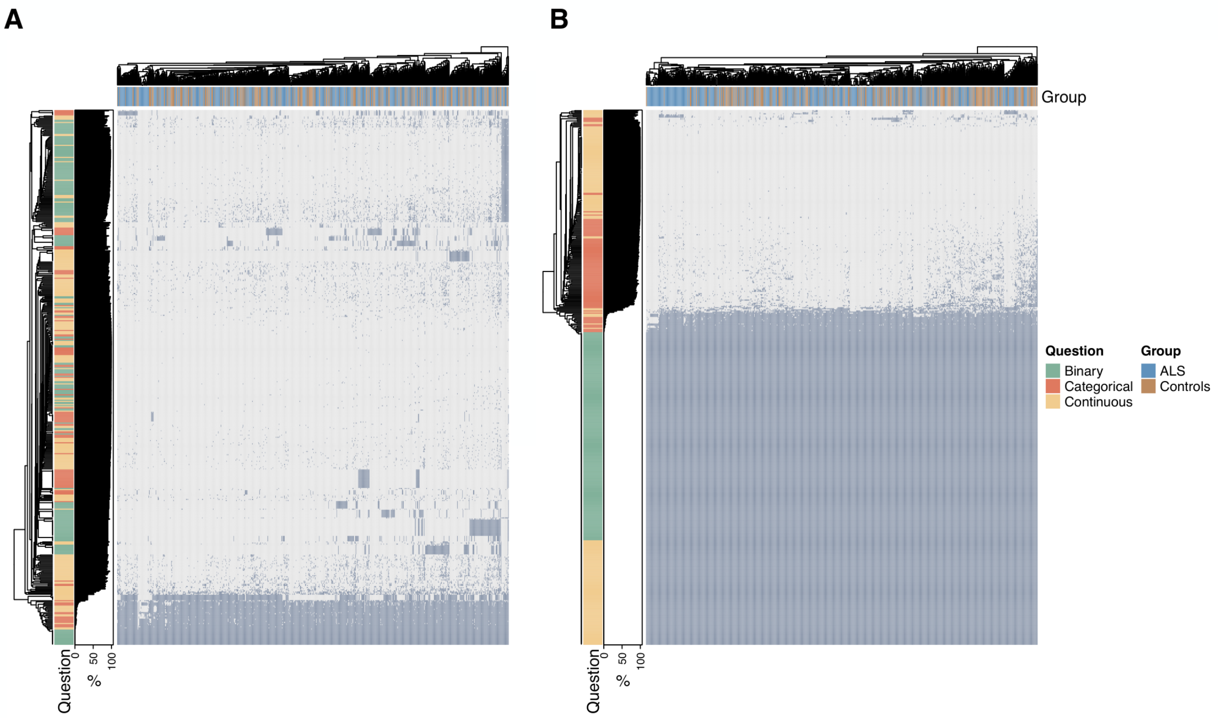


**Supplementary Figure 1: Heatmaps before and after manual imputation.**
**(A)** Overview of responses to the original questionnaire. Dark grey cells represent provided answers, where white cells indicate questions that were left unanswered. Participants were instructed to leave items blank if they were not applicable or if they were unsure, resulting in a considerable number of non-responses in the initial dataset. **(B)** Overview after manual imputation. For selected binary items and questions about the number of visits to specific healthcare specialists (continuous), missing responses were interpreted as "no" or zero visits, under the assumption that non-response indicated non-occurrence during the specified time frame. In both panels, questions are categorized as binary, categorical or continuous, and the percentage of responses (i.e., answer rate; %) is shown on the left-hand side of each heatmap. Group affiliation (ALS or controls) is indicated in the upper annotation of each heatmap.

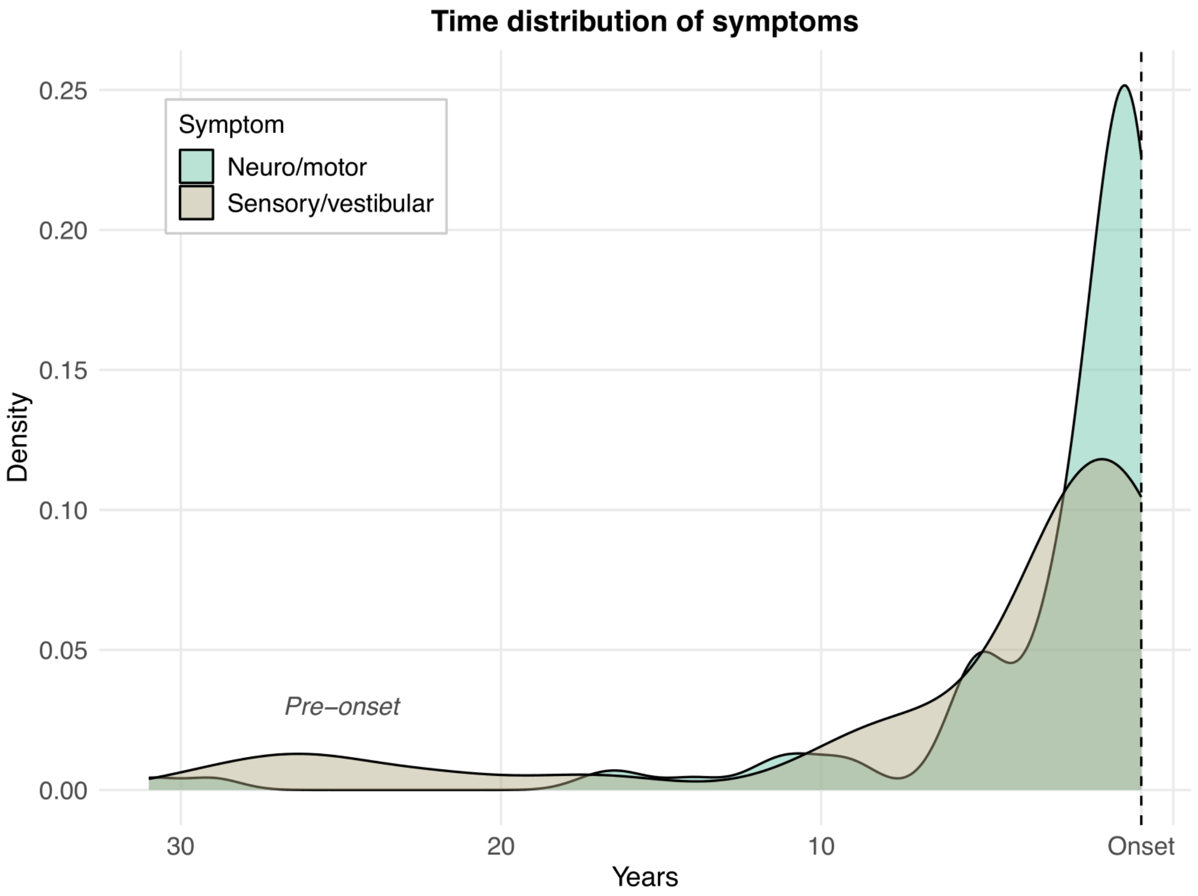


**Supplementary Figure 2: Time distribution of neuro/motor and sensory/vestibular symptoms in ALS patients.**

Density distributions of reported neuro/motor and sensory/vestibular symptoms, quantified in years relative to the date of ALS symptom onset (Onset). Only responses for symptoms spanning the pre-onset period were analysed and visualized. The density of responses is coloured by symptom type.

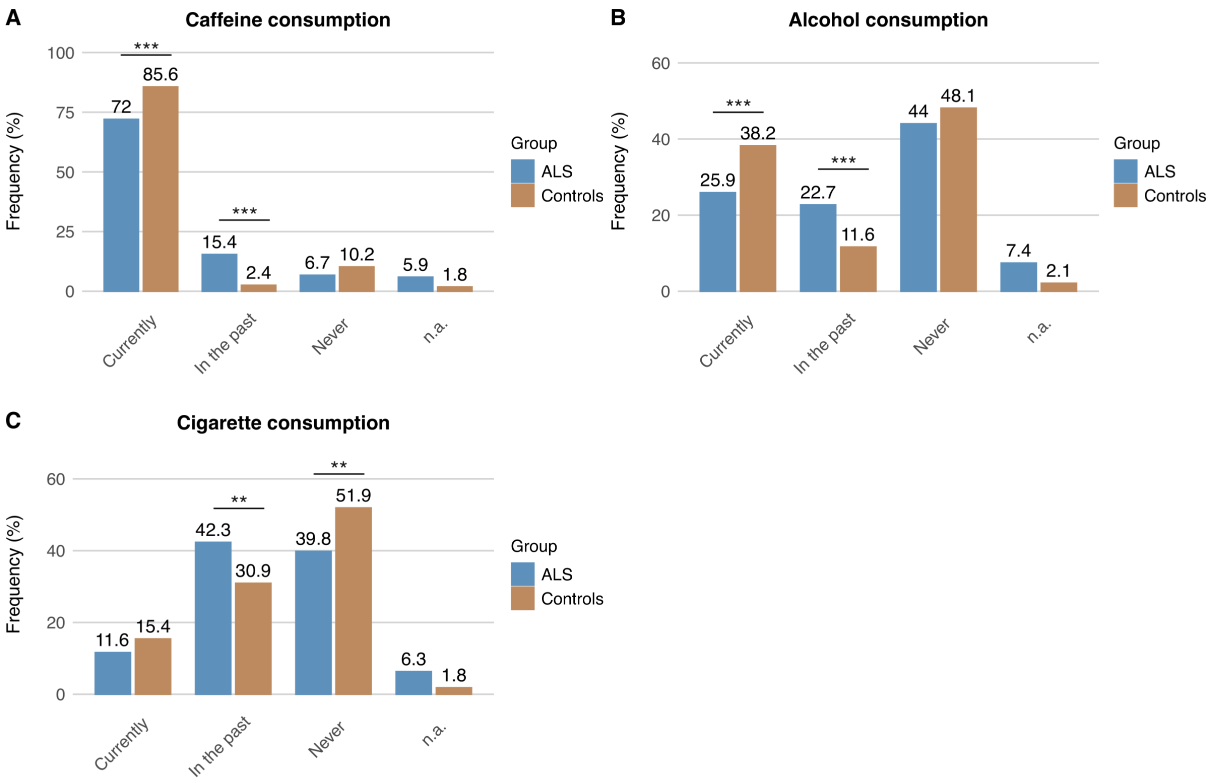


**Supplementary Figure 3: Substance use among ALS patients and control participants.** Bar graphs displaying patterns of substance use, including **(A)** caffeine, **(B)** alcohol, and **(C)** cigarette consumption, categorized as current use, past use, or never. Participants could select only one response category for each substance. Missing values for substance consumptions were categorized as not available (n.a.). Significance was assessed using univariate logistic regression analyses stratified by sex and is indicated as $P < 0.05$ (*), $P < 0.01$ (**), $P < 0.001$ (***).

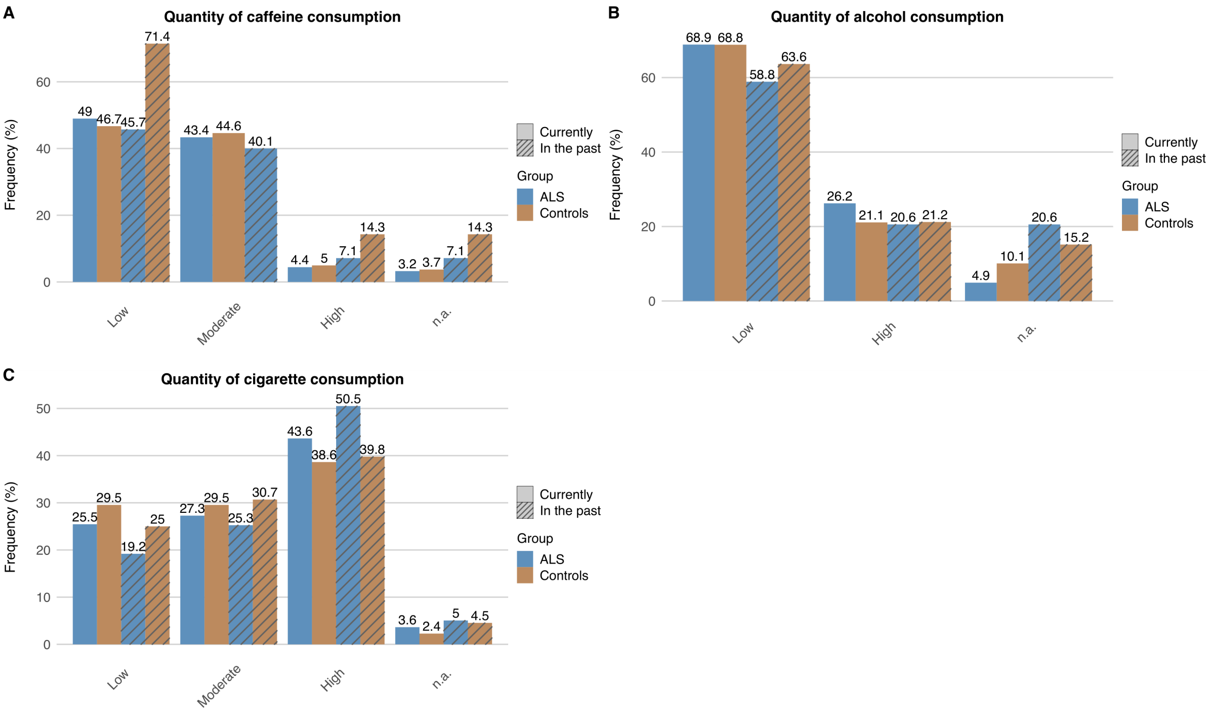


**Supplementary Figure 4: Quantity of substance use among ALS patients and control participants.**

Bar graphs illustrate the quantities of substances, such as **(A)** caffeine, **(B)** alcohol, and **(C)** cigarette consumption. Only participants who reported current or past consumption of each substance were included in these distributions. For caffeine and alcohol, participants reported their typical intake based on predefined consumption categories. Specifically, **(A)** Caffeine consumption was classified as low (1–2 cups of coffee/day), moderate (3–5 cups/day), and high (>5 cups/day). Participants were instructed that one cup of black coffee corresponds approximately to the caffeine content of one liter of cola or one can of an energy drink. **(B)** Alcohol consumption was classified based on typical daily intake, with a standard drink defined as 0.25 L beer, 0.1 L wine or sparkling wine, or 0.04 L spirits; low consumption was ≤1 drink/day for women or ≤2 drinks/day for men (≤5 days/week), and high consumption was above these thresholds. **(C)** For smoking, participants indicated the number of cigarettes smoked per day. To standardize reporting, smoking quantities were grouped into three categories: low (1–6 cigarettes/day), moderate (7–14 cigarettes/day), and high (>14 cigarettes/day), based on the reported amounts. The frequencies of substance consumption were calculated based on all participants who reported current or past use, respectively. Missing values for substance quantities were categorized as not available (n.a.).

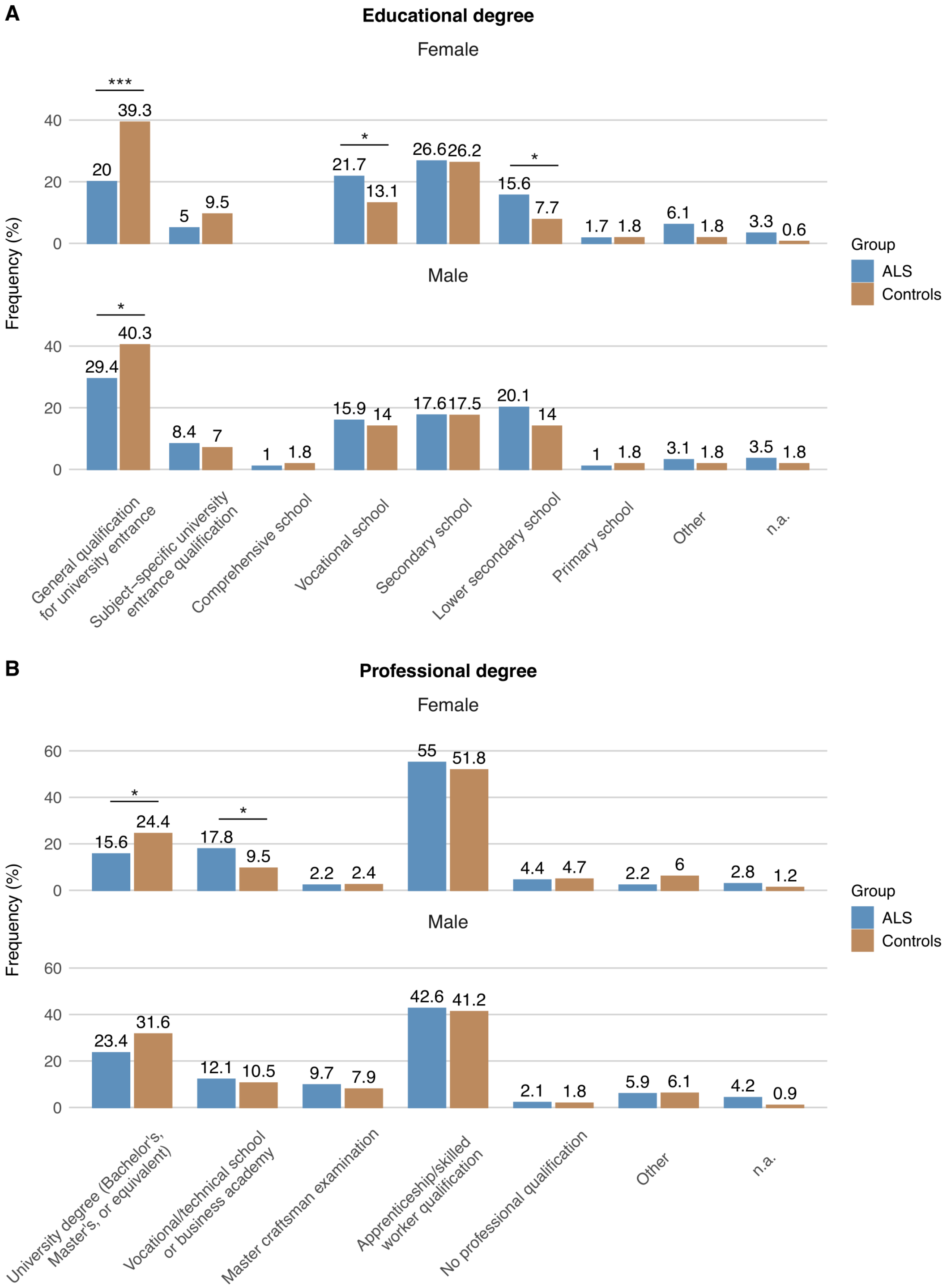


**Supplementary Figure 5: Highest educational and professional qualifications among ALS patients and control participants, stratified by sex.**
Bar graphs show the distribution of **(A)** educational degrees and **(B)** professional qualifications for female and male participants separately.
Missing educational and professional qualifications of participants were categorized as not available (n.a.). Significance was assessed using univariate logistic regression analyses stratified by sex and is indicated as $P < 0.05$ (*), $P < 0.01$ (**), $P < 0.001$ (***).

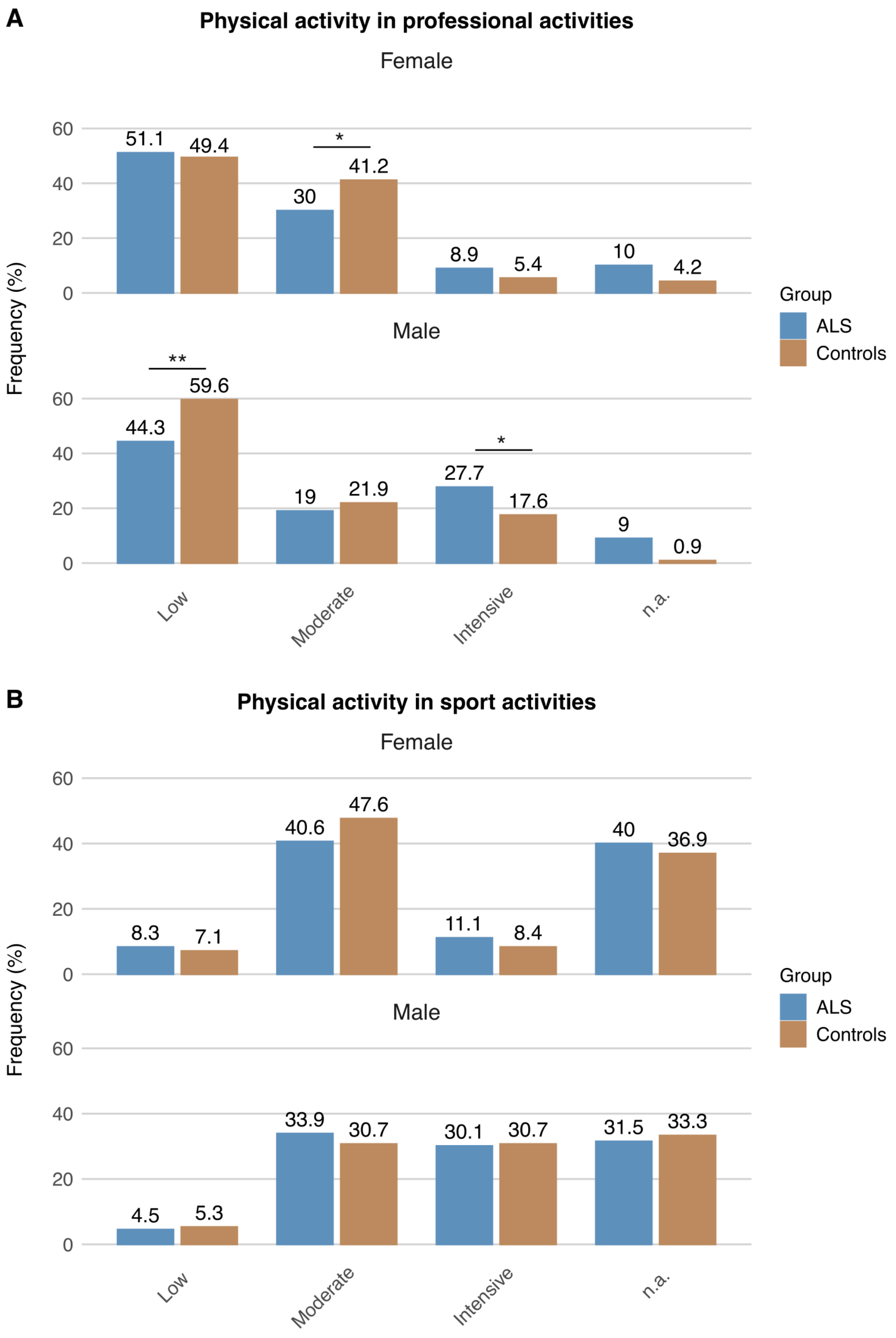


**Supplementary Figure 6: Occupational physical activity and regular sports participation among ALS patients and control participants, stratified by sex.**

Bar graphs displaying the distribution of **(A)** physical activity related to occupation, categorized as low, moderate, or high intensity, and **(B)** regular sports participation, also classified into low, moderate, or high intensity, presented separately for male and female participants. Missing physical activity in professional and sport activities of participants were categorized as not available (n.a.). Significance was assessed using univariate logistic regression analyses stratified by sex and is indicated as $P < 0.05$ (*), $P < 0.01$ (**), $P < 0.001$ (***).

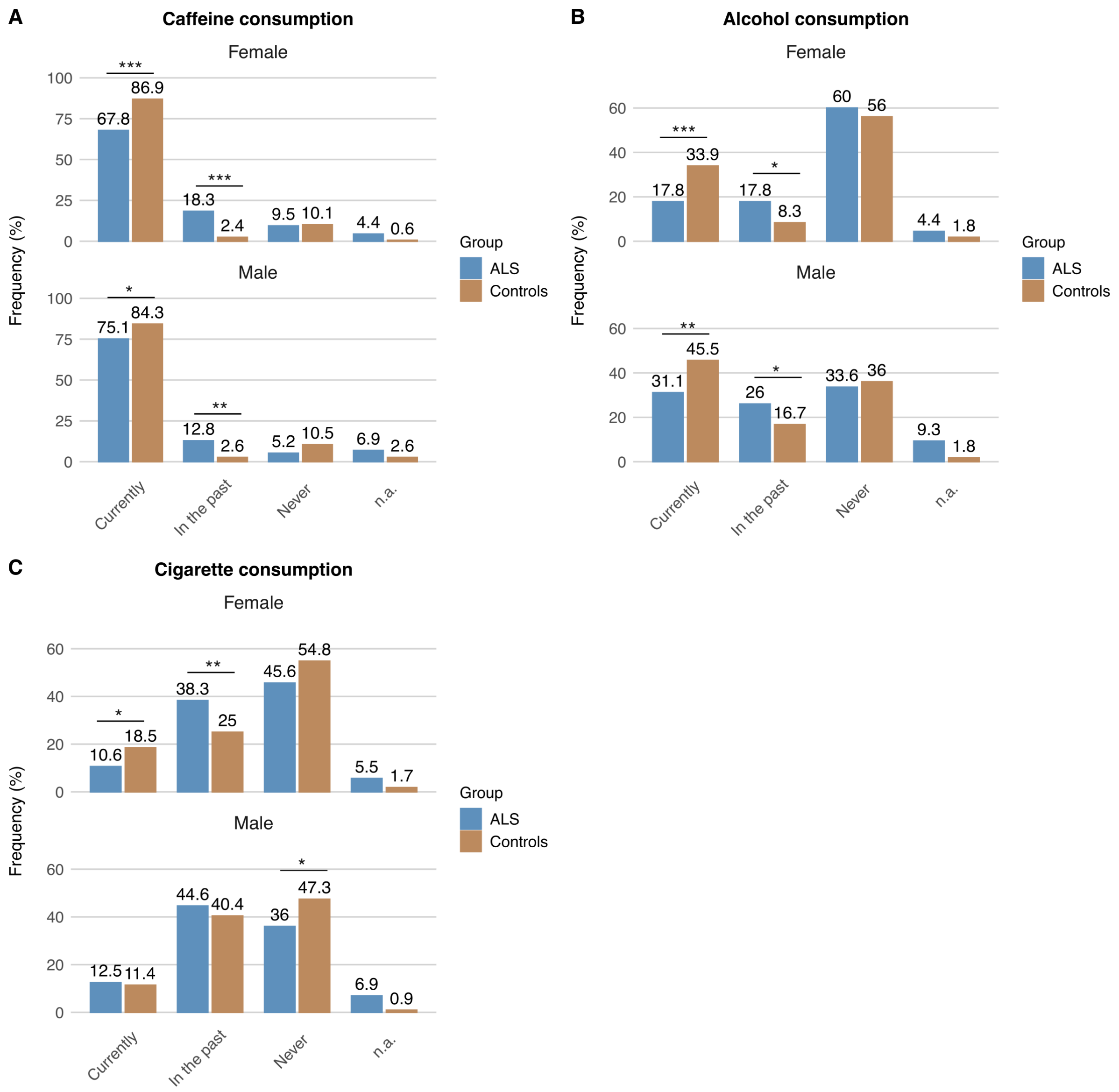


**Supplementary Figure 7: Substance use among ALS patients and control participants, stratified by sex.**

Bar graphs displaying patterns of substance use, including **(A)** caffeine, **(B)** alcohol, and **(C)** cigarette consumption, categorized as current use, past use, or never. Data are presented separately for female and male participants.

Missing substance consumptions of participants were categorized as not available (n.a.). Significance was assessed using univariate logistic regression analyses stratified by sex and is indicated as $P < 0.05$ (*), $P < 0.01$ (**), $P < 0.001$ (***).
