## Supplementary material for "EARLY-ALS: A Multicentre Study on Presymptomatic and Prodromal Amyotrophic Lateral Sclerosis": Table 1

**Table 1 Study group characteristics of patients with ALS and controls**

|  | **ALS**  (n=475) | **Controls**  (n=285) | ***P*** |
| --- | --- | --- | --- |
| **Demographics** |  |  |  |
| Age [years], median (IQR)  n.a., n (%) | 63 (55, 70)  233 (49.05) | 61 (53, 67)  164 (57.54) | 0.073^a^ |
| Sex, n (%)  Female  Male  n.a. | 180 (37.90)  289 (60.84)  6 (1.26) | 168 (58.95)  114 (40.00)  3 (1.05) | < 0.001^b^ |
| **Composition of control group**  Relationship to proband, n (%)  Parents  Siblings  Children  Uncle  Partners  Other  n.a.  **Disease characteristics** |  | 20 (7.02)  10 (3.51)  31 (10.88)  1 (0.35)  214 (75.09)  6 (2.10)  3 (1.05) |  |
| Age at onset [years], median (IQR)  n.a., n (%) | 60 (51.8, 68)  247 (52.00) | - |  |
| Site of onset, n (%)  Spinal  Bulbar  n.a.  ALSFRS-R, median (IQR)  Known pathogenic variant, n (%)  *C9orf72*  *SOD1*  *TARDBP*  *ANXA11*  *TBK1*  *SETX*  n.a.    ALS subtype, n (%)  Classical ALS (Charcot-type)  Progressive bulbar palsy (PBP)  Flail-arm syndrome  Primary lateral sclerosis (PLS)  Flail-leg syndrome  Progressive muscular atrophy (PMA)  ALS-FTD  n.a. | 378 (79.58)  75 (15.79)  22 (4.63)  33 (25, 38)  9 (1.89)  4 (0.84)  1 (0.21)  1 (0.21)  1 (0.21)  1 (0.21)  458 (96.42)  330 (69.47)  39 (8.21)  22 (4.63)  22 (4.63)  17 (3.58)  13 (2.74)  4 (0.84)  28 (5.90) | -  -  -  -  -  -  -  -  -  -  -  -  -  -  -  - |  |

Continuous variables are presented as median and interquartile range (IQR), and categorical variables are reported as absolute numbers and percentages. Within the control group, the “Other” category included friends, caregivers, stepchildren and sons- or daughters-in-law.

ALS, Amyotrophic lateral sclerosis; ALSFRS-R, Amyotrophic Lateral Sclerosis Functional Rating Scale – Revised; FTD, Frontotemporal dementia, n.a., not available.

^a^ Mann-Whitney test.

^b^ Fisher’s exact test.
